# Simulating tDCS-induced electric fields in stroke patients: realistic-lesion head models are needed

**DOI:** 10.1101/2025.09.23.25336427

**Authors:** Ikko Kimura, Marcus Meinzer, Daria Antonenko, Robert Darkow, Agnes Flöel, Axel Thielscher

## Abstract

**Introduction:** Transcranial direct current stimulation (tDCS) is tested as tool for post-stroke rehabilitation in aphasia, and individualized simulations of tDCS-induced electric fields (E-fields) can guide its application. However, the accuracy of simulations is challenged by complex and variable tissue properties of stroke lesions. Here, we assessed the impact of stroke lesions on tDCS-induced E-fields realistically in terms of lesion size, shape, and conductivity.

**Methods:** Structural and diffusion MRI datasets of stroke patients with aphasia (n=13, six females, age=38-70 years) and age-matched healthy controls (n=13, eight females, age=24-76 years) from a previous study were analyzed. Simulated E-fields were first compared between healthy head models with and without artificial lesions homogenously filled with cerebrospinal fluid. Then, the effects of lesion heterogeneity were tested by comparing E-fields for models of stroke patients with homogenous versus inhomogeneous (realistic) lesion conductivity informed by diffusion-to-conductivity mapping.

**Results:** Adding artificial lesions to healthy head models altered the magnitude of E-fields (|E|) near the target region (ROI) by up to 47%. Diffusion-to-conductivity mapping revealed substantial variability in lesion conductivities within and across patients. Modifying homogenous to realistic lesion models showed small to moderate |E| differences within the ROI depending on montage type, lesion size, and lesion-to-target distance.

**Conclusion:** Stroke lesions affect tDCS-induced E-fields with substantial variability across montages and individuals. These findings support the use of head models that include realistic representations of the shape, size and conductivity of the lesions to improve the accuracy of individualized tDCS simulations and guide personalized stimulation protocols in stroke rehabilitation.

## 1. Introduction

Stroke is the most frequent cause of acquired impairment of language and communication (aphasia) and ∼20-40% of patients develop chronic symptoms, requiring long-term use of rehabilitation services (El Hachioui et al., 2013; Pedersen et al., 2004). Intensive and individually tailored speech-language therapy (SLT) can improve chronic aphasia (Brady et al., 2025, 2016), but recent clinical trials have also highlighted variable outcomes across patients and high percentages of non-responders (Breitenstein et al., 2017; Palmer et al., 2019; Rose et al., 2022). This has generated interest in adjunctive treatments capable of enhancing neuroplastic processes in aphasia, thereby rendering the lesioned brain more receptive to behavioral interventions (Crosson et al., 2019).

Based on studies in healthy individuals demonstrating that transcranial direct current stimulation (tDCS) can enhance language processing and learning (e.g., Flöel et al., 2008; Martin et al., 2017; Meinzer et al., 2014; Perceval et al., 2020, 2017), this approach has received substantial attention in aphasia research (Flöel et al., 2011; Meinzer et al., 2016; Brady et al., 2025; Elsner et al., 2019; Fridriksson et al., 2018). TDCS uses two or more scalp-attached electrodes to administer a weak electrical current to modulate excitability and neuroplasticity in target brain networks (Stagg and Nitsche, 2011). It is easy to administer and offers an effective placebo mode and an excellent safety profile, even in stroke patients (Antal et al., 2017). These features make tDCS an attractive tool for experimental or clinical studies in aphasia.

To date, dozens of aphasia trials that combined a wide variety of SLT and tDCS approaches have been completed (Meinzer et al., 2025). However, meta-analytic evidence for significant enhancement of SLT effects by tDCS is mixed and positive results have mainly been shown for specific linguistic functions like picture naming (Brady et al., 2025; Elsner et al., 2019; Raymer and Johnson, 2024), rather than functional communication abilities. Hence, a more personalized approach to using tDCS in post-stroke aphasia may be necessary.

For example, targeting of specific brain regions with tDCS can be aided by individualized computer simulations that estimate the distribution and intensity of the induced electrical field (E-field) using structural magnetic resonance imaging (MRI) data (Datta et al., 2011; Evans et al., 2023; Galletta et al., 2015; Krishnamurthy et al., 2025; Richardson et al., 2015; Minjoli et al., 2017). In treatment planning, these simulations can be valuable to ensure that the induced E-field is delivered to the intended target regions for tDCS and at a sufficient dose. However, the quality of E-field simulations critically depends on the validity of assumptions about the conductive properties of different tissue classes in the respective head models (Hunold et al., 2023). In stroke patients, this is further complicated by complex lesion shapes and sizes as well as variable tissue properties of the lesioned areas (Duering et al., 2020), that have not or only partly been accounted for in most previous studies (e.g., the lesion was classified as cerebrospinal fluid [CSF]; Evans et al., (2023); but see Krishnamurthy et al., (2021)).

In the present study, we used datasets of patients with post-stroke aphasia and age-matched healthy controls from a previous study (Darkow et al., 2017) to examine how stroke lesions influence tDCS-induced E-fields. Specifically, this study is comprised of two comparisons: (1) simulated E-fields from healthy head models with and without artificial homogeneous lesions derived from the stroke patients, and (2) those from head models of stroke patients with and without considering spatial heterogeneity of lesion conductivity. This study has three unique features. First, artificial lesions applied in this study are realistic in size and shape, in contrast to simplified lesion shapes applied in some of the previous studies (e.g., Evans et al., 2023). Second, we estimated the inter- and intra-individual variability of lesion conductivity using diffusion-to-conductivity mapping (Tuch et al., 2001). Third, we evaluated potential effects using focal montages (Niemann et al., 2024) as well as conventional bipolar montages.

## 2. Methods

### 2.1. Participants

We used an existing dataset of 16 stroke patients and a group of 16 age-matched healthy controls, which performed tDCS over hand representation of the left primary motor cortex (M1) to assess the brain activity changes during a picture naming task (Darkow et al., 2017). In this dataset, T1-weighted (T1w) and diffusion MRI (dMRI) data were obtained for chronic stroke patients and healthy controls. Three pairs of stroke patients and individually matched healthy controls were excluded because dMRI data were lacking in the patients (two pairs), or due to coregistration issues between dMRI and T1w data (one pair). This resulted in 13 pairs of stroke patients (six females; age: mean = 55.5 years, standard deviation [SD] = 10.0 years, 38-70 years; time since stroke: mean = 52.8 months, SD = 49.4 months, 12-169 months) and age-matched healthy controls (eight females; age: mean = 55.3 years, SD = 18.6 years, 24-76 years). A two-tailed paired t-test confirmed that there were no statistical differences in age between the 13 included stroke patients and the healthy controls (t_12_ = −0.02, *P* = 0.98).

### 2.2. Image acquisition

All T1w and dMRI data were collected using a Siemens Trio 3T scanner equipped with a 16-channel array head coil (Siemens, Erlangen, Germany) in the Berlin Centre for Advanced Imaging of the Charité University Hospital. Detailed imaging protocols can be found in Darkow et al., (2017) for T1w. Briefly, T1w data were obtained with an MPRAGE sequence (voxel size = 1 × 1 × 1 mm³, echo time [TE] = 2.52 msec, repetition time [TR] = 1900 msec, inversion time = 900 msec, flip angle = 9 degree). DMRI were recorded with a two-dimensional single-shot spin-echo echo-planar sequence (voxel size = 2.3 × 2.3 × 2.3 mm³, matrix size = 96 × 96 × 61, b-value = 1000 s/mm^2^, number of directions = 64, TR = 7500 ms, and TE = 86 ms).

### 2.3. Image preprocessing and generation of head models

T1w data were automatically segmented using *charm* implemented in SimNIBS (ver 4.1.0; Puonti et al., (2020)). This method segments the whole head into eight tissues (gray matter [GM], white matter [WM], CSF, compact bone, spongy bone, scalp, large blood vessels, eye muscles, and eye balls) to create a tetrahedral mesh representing the head anatomy (the “head model”). The CSF was further divided into ventricular and cortical CSF following Gregersen et al. (2024) to separate the ventricular CSF voxels from those that may contain meninges or were inside the lesion. The ventricular CSF was subsequently used to calibrate the diffusion-to-conductivity mappings in the stroke patients, as detailed in *2.5. Modeling of lesion conductivities using dMRI data*. For stroke patients, lesion masks from the previous study (Darkow et al., 2017), where the outlines of lesions were manually delineated by T1w and fluid attenuation inversion recovery (FLAIR) images, were added to the head model using the *add_tissues_to_upsampled* function in SimNIBS. These lesion masks were homogeneously filled with CSF (homogeneous-lesion head models).

DMRI data were preprocessed using *dwi2cond* from SimNIBS. This command mainly utilizes tools from the FMRIB Software Library (FSL; ver 6.0.4) (see Mosayebi-Samani et al., (2025) for details). In brief, dMRI data were eddy-current corrected, and co-registered to the T1w data. Here, distortion correction was not performed because images required for that process (non-diffusion weighted images with inverse phase encoding direction or field map) were not acquired in this dataset. Instead, dMRI data were non-linearly co-registered to the T1w data to mitigate the effect of B0 inhomogeneity-induced distortions using FNIRT with custom parameters.

### 2.4. Modification of healthy head models to assess effect of lesions on the E-field

To evaluate the effect of lesions on the E-field, artificial lesions were added to the head models of the healthy control group (i.e., healthy head models vs. homogeneous-lesion head models; Fig. 1A). To achieve realistic lesion shapes, the added lesion masks were created from the matched pair of stroke patients. Specifically, the lesion masks of the stroke patients were non-linearly transformed to the space of their matched healthy controls by first warping them to MNI space using *subject2mni* from SimNIBS, and then further to the matched healthy controls space using *mni2subject* in SimNIBS. Slight overlaps of the transformed lesion masks with the skull or scalp were excluded. The final masks were added to the head models of the healthy controls using the same approach as for the homogeneous-lesion head models of the stroke patients (see *2.3. Image preprocessing and generation of head models* for details).

**Figure 1.**
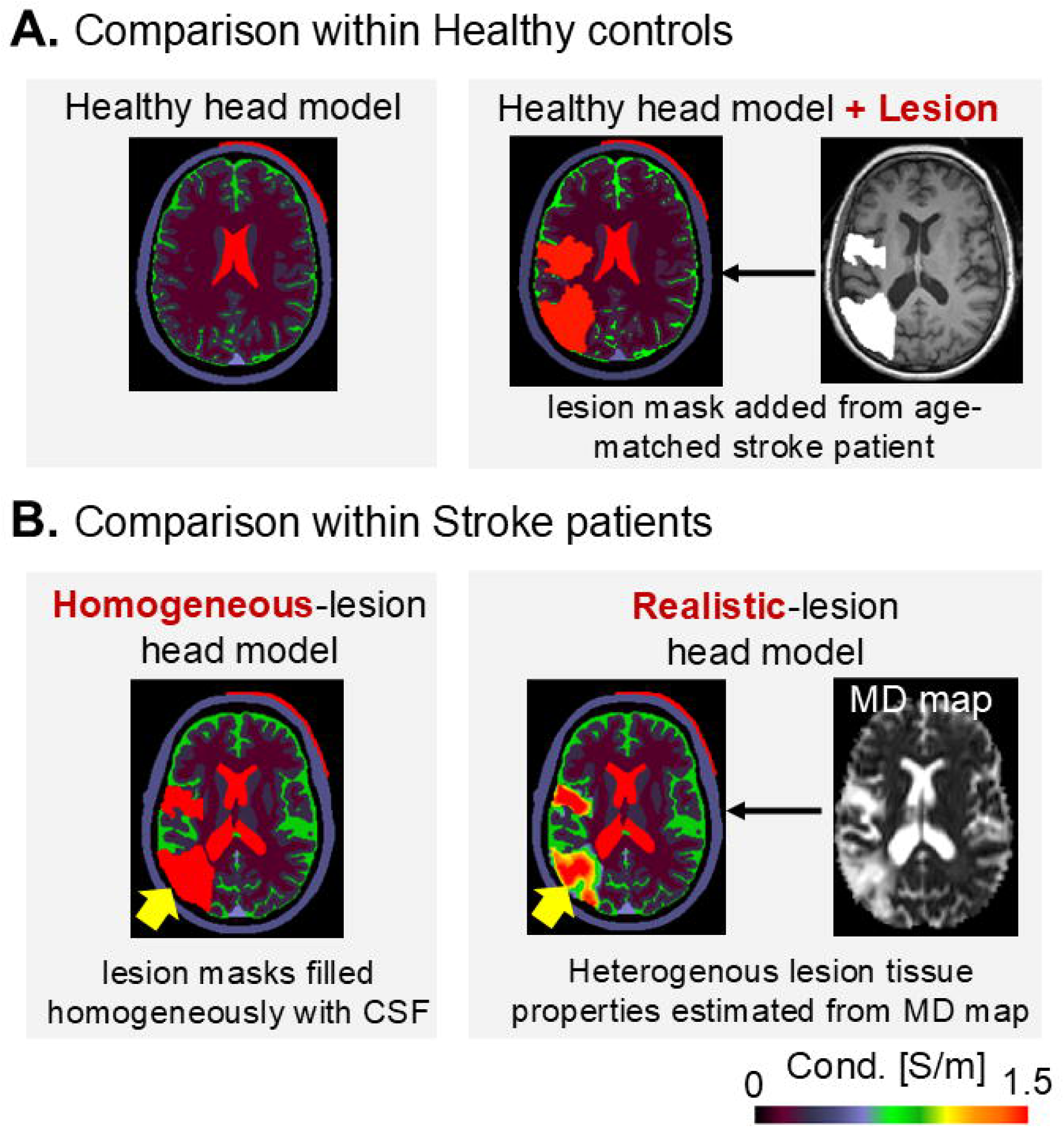
Head models compared in this study. This study consists of two comparisons of simulated electric fields (E-fields): (A) Within head models of age-matched healthy controls of stroke patients, simulations were performed without (healthy head model) and with (healthy head model + lesion) artificially included lesions. To ensure realistic lesion shapes and sizes, we created the lesion masks by non-linearly transforming them from the age-matched stroke patients. (B) Within stroke patients, the simulated E-field was compared between head models with homogeneously (homogeneous-lesion head model) and realistically (realistic-lesion head model) modeled lesions (yellow arrows). The “homogeneous-lesion” head models set the conductivity of lesions to the conductivity of cerebrospinal fluid (CSF; left panel), while the “realistic-lesion” models estimated the conductivity within lesions from mean diffusivity (MD) maps derived from diffusion MRI. The figure in each panel represents the set conductivity for each head model.

### 2.5. Modeling of lesion conductivities using dMRI data

The conductivity of lesions is usually modelled as homogeneous CSF. However, this is an over-simplification, as lesions usually consist of a mixture of CSF and various tissues, including scar tissue. To estimate the impact of this simplification on the simulated E-field, additional head models were created for the stroke patients. For these “realistic-lesion head models” (Fig. 1B), the spatially inhomogeneous and isotropic conductivities within the lesions were estimated from the dMRI data (diffusion-to-conductivity mapping). This was achieved by applying a linear relationship between the ohmic conductivity and the diffusivity of water, as measured by dMRI (Tuch et al., 2001):

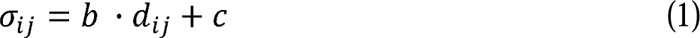

Here, σ*_ij_* and *d_ij_* denote the *j*-th (j∈{1,2,3}) ohmic conductivity and diffusivity eigenvalues at voxel *i*, respectively. We assumed the conductivities within lesion masks to be isotropic, and hence estimated average conductivities across directions as follows:

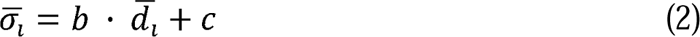

*σ̅_i_* and *d̅_i_* represent the mean ohmic conductivity and the mean diffusivity (MD) at voxel *i*, respectively. As we presumed the conductivity of lesions to be between the conductivity of GM and CSF, the slope (*b*) and intercept (*c*) were estimated for each patient by fitting the literature values of conductivities of GM (0.275 S/m) and ventricular CSF (1.79 S/m) against their MD values (Supp. Fig. 8). We restricted CSF to the ventricles, because modeling studies suggested that thin cortical CSF area might be confounded with meninges (Jiang et al., 2020). Visual inspection of the fitted conductivities confirmed that they were in the expected range also for WM, which was left out from the fitting as control tissue (Supp. Fig. 8). The conductivity of cortical CSF was estimated to lie between that of GM and ventricular CSF. Finally, the conductivities within the lesions were estimated from the dMRI data using the fitted model, setting the upper and lower limit to be 1.79 S/m and 0.008 S/m (SimNIBS’s default value for ventricular CSF and compact bone), respectively.

### 2.6. E-field Simulations

Four electrode montages were used to simulate the E-fields induced by each montage of tDCS, comprising two target locations [M1 or a personalized peri-lesional target derived from functional MRI], each with two types of montages [bipolar or focal montage] (Figs. 2A-D). The peri-lesional target was defined for each stroke patient as the location of the peak activation closest to the lesion, determined via functional MRI of a picture naming task in the previous study (Darkow et al., 2017; Fig. 2E). This approach has widely been used in aphasia patients to target individually determined functional perilesional cortex (Baker et al., 2010; Fridriksson et al., 2018). The peri-lesional target locations were non-linearly transformed to the corresponding paired healthy controls. One participant was excluded for the montages of the peri-lesional target as there was no peak activated location near the lesions. Consequently, 12 pairs of stroke patients and their matched healthy controls were further analyzed for the analyses related to the peri-lesional target stimulation.

**Figure 2.**
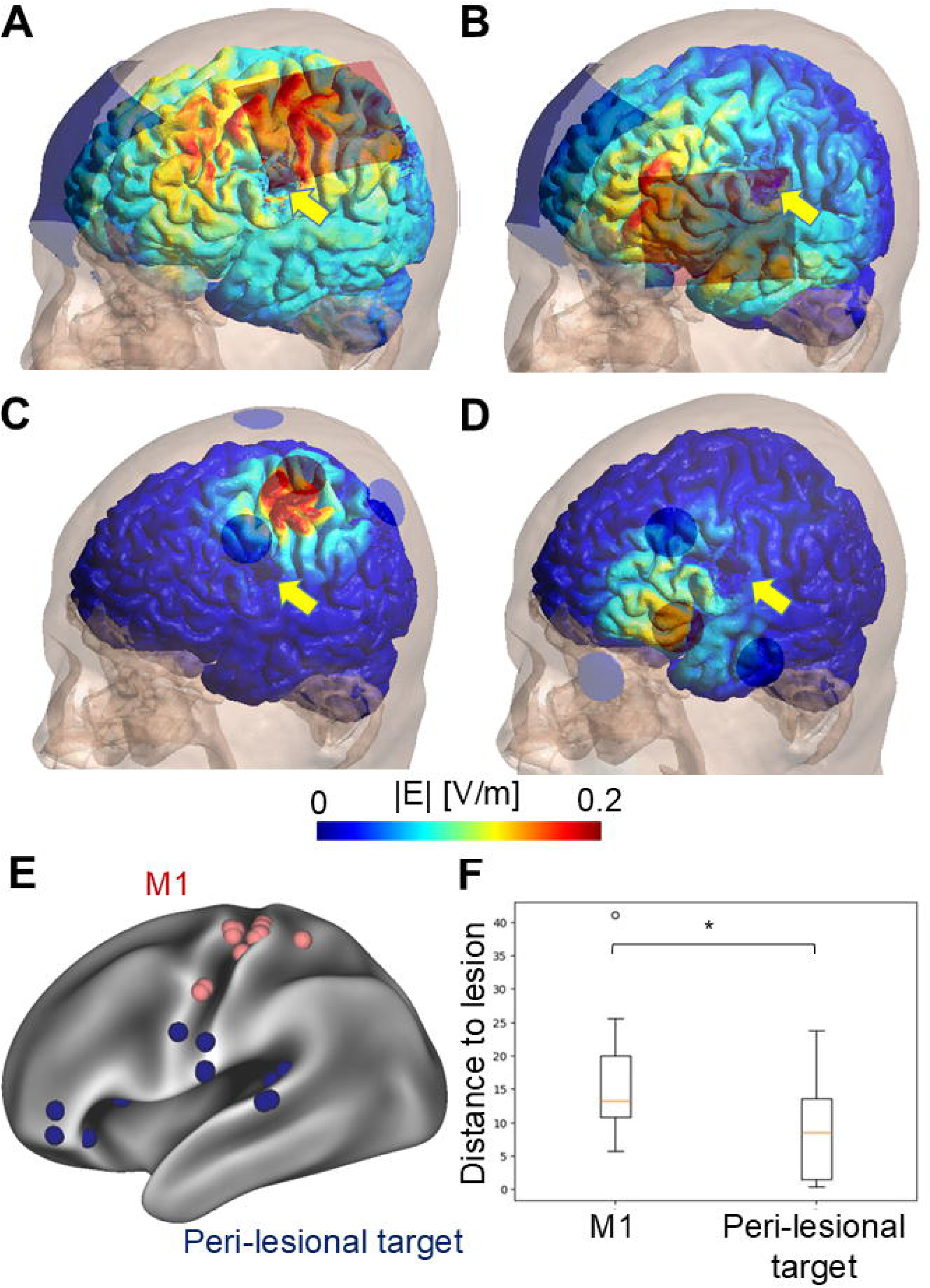
Simulated montages. (A-D) represent the simulated electric fields (E-fields) on the gray matter in a representative stroke patient for the four tDCS montages applied in this study. We simulated the E-fields on bipolar montages targeting the hand representation of left primary motor cortex (M1; A) or peri-lesional target (B), as well as focal montages placed above M1 (C) or the peri-lesional target (D). The red and blue electrodes indicate anodes and cathodes, respectively, and yellow arrows denote the lesion. (E) The M1 (red) and peri-lesional target locations (blue) for each stroke patient on the fsaverage_LR32k template. (F) Boxplot of the closest distance between the lesions and the M1 or peri-lesional targets. * *P* < 0.05

For the bipolar montages, the anode was located over the target location, while the cathode was placed on the right supraorbital area (just above the right eyebrow) using rectangular rubber electrodes (anode: 5 × 7 cm^2^, cathode: 10 × 8 cm^2^; Figs. 2A and B). For the focal montages, the anode was positioned over the target location, while the three cathodes were equally distributed with a radius 4.5 cm using the SimNIBS function *tDCS_Nx1.py* (Niemann et al., 2024; Figs. 2C and D). One of the cathodes in the focal montage was placed in the direction of C3 to C1 for M1, while it was in the direction of left pre-auricular to T7 for the peri-lesional target. All rubber electrodes used in the focal montage were assumed to be circular with a radius of 12.5 mm. For M1 stimulation, the anode was located at C3 for the bipolar montages following Darkow et al. (2017), while it was placed over the left hand-knob for the focal montages (Niemann et al., 2024). For peri-lesional target stimulation, the anode was centered over the individual peri-lesional target for both the bipolar and focal montages.

The finite element method implemented in SimNIBS was used to simulate the E-field induced by each montage of tDCS (Thielscher et al., 2015). All rubber electrodes were assumed to be two mm thick with one mm of electrode gel. The default conductivity values in SimNIBS were applied for the simulation, unless otherwise noted. The conductivity was set to 0.85 S/m for cortical CSF (Jiang et al., 2020) and 1.79 S/m for ventricular CSF (Baumann et al., 1997). In the homogeneous-lesion head model for the healthy controls and stroke patients, the lesion conductivity was fixed to 1.79 S/m, which corresponds to the conductivity of ventricular CSF as described above. For the bipolar montages, the stimulation currents injected into the electrodes were set to ±1 mA. For the focal montages, the current injected into the anode was set to 1 mA and that of each cathode to −1/3 mA. The magnitude (|E|) and normal component (nE) of the E-field (Antonenko et al., 2019) were extracted at the middle layer of the cerebral cortices (i.e., the midpoint of pial and white surfaces; cortical middle layer) of both hemispheres for further analyses. For healthy controls, the cortical surface areas corresponding to the artificially added lesions were excluded.

Differences in the simulated E-fields were assessed between (1) head models of healthy controls with versus without artificially added lesions (Fig. 1A), or (2) head models of patients with homogenously versus realistically modelled lesions (Fig. 1B). For both cases, E-fields were simulated for the bipolar and focal montages and compared. For analysis, global (across overall grey matter) and local (nearby the target location) metrics of the E-field differences at the cortical middle layer were extracted. For the global metrics, the peak value and focality of |E| within the cerebral cortex were calculated. Using SimNIBS’s default settings, the peak value was defined as the 95th percentile of |E|, while the focality was defined as the area where |E| exceeds 75% of the 95th percentile of |E|. For the local metrics, |E| or nE at the cortical middle layer was extracted within region-of-interests (ROIs). The ROIs were defined for each montage as a 12.5 mm radius sphere centred at the target location, with the size matched to the size of the electrodes in the focal montages (Niemann et al., 2024). The following measures were used for assessing the E-field differences between the head models within the ROI:

i. the differences in the mean |E| or nE (Δmean |E| or nE)

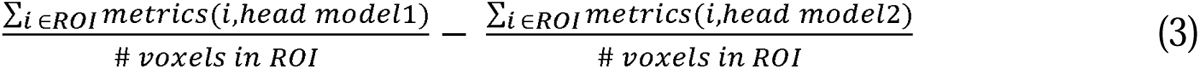
ii. the mean of the position-wise absolute differences in |E| or nE (mean |Δ|E|| or |ΔnE|)

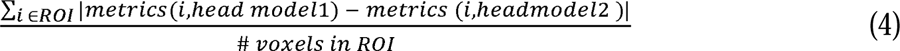

Here, head model 1 is the healthy head model with artificial lesions or realistic-lesion head model; head model 2 is the healthy head model or homogeneous-lesion head model; and *metrics* are Δmean |E|, Δmean nE, mean |Δ|E|| or mean |ΔnE|. The first measures, Δmean |E| or nE (eq. 3), were used to assess the overall increase or decrease in E-field within the ROI, irrespective of more subtle changes of the spatial stimulation patterns within the ROI. The second measures, mean |Δ|E|| or |ΔnE| (eq. 4), were used to detect more local changes of the spatial stimulation patterns within the ROI (if the lesion increased the E-field in some locations and decreased in others within the ROI, these differences would cancel each other out in Δmean |E| or nE, but not in mean |Δ|E|| or |ΔnE|). For the both measures, the relative differences compared to the healthy head model or the homogeneous-lesion head model (Evans et al., 2023) were additionally calculated.

### 2.7. Statistical analysis

Two-tailed one-sample t-tests were used to detect consistent increases or decreases in the global and local difference measures across participants. Two-tailed two-sample t-tests were applied to evaluate whether the measures or differences in the measures were different across bipolar and focal montages for each target location (M1 or peri-lesional target). As the local difference metrics showed high inter-individual variability in both stroke patients and healthy controls (see Results), Spearman’s correlation coefficients were additionally calculated between the inter-individual variations in the difference in local metrics within the ROI and anatomical lesion features. The following lesion features were used: (1) lesion size in mm³, (2) lesion-to-target distance in mm (i.e., the shortest distance between a target and lesion masks), and (3) mean absolute conductivity difference within lesion masks in S/m. As these are exploratory analyses, *P* < 0.05 uncorrected for multiple comparisons was set as threshold for statistical significance. Statistical functions in SciPy (ver 1.10.1) and JASP (ver. 0.18.3 for Windows) were used for all statistical analysis.

## 3. Results

### 3.1. Differences in E-fields between head models with and without lesions

Compared to the stroke patients, the artificially created lesions in the age-matched healthy controls were not significantly different in the size (t_12_ = −0.52, *P* = 0.61; Supp. Fig. 5A) or the distances to the left M1 (t_12_ = −1.15, *P* = 0.27; Supp. Fig. 5B) or peri-lesional target (t_11_ = −1.15, *P* = 0.15; Supp. Fig. 5C). This suggests that the main lesion features were maintained even after transforming the lesions from the stroke patients to the age-matched healthy controls. Between the head models with and without artificial lesions, the mean absolute difference in conductivity within the region of the artificial lesions showed small variability across the participants (mean = 1.55 S/m, SD = 0.05 S/m, 1.48-1.63 S/m).

#### Global metrics

Adding the lesions to the healthy head models changed the 95% peak |E| by up to 15% (mean = 7.8%, SD = 4.4%), 27% (mean = 8.2%, SD = 7.6%), 34% (mean = 9.0%, SD = 8.8%), and 31% (mean = 18%, SD = 7.5%) for the bipolar M1, focal M1, bipolar peri-lesional target, and focal peri-lesional target montages, respectively (Supp. Fig. 6C). In most participants, adding lesions reduced the 95% peak |E| (bipolar M1: t_12_ = −3.18, *P* = 0.008; focal M1: t_12_ = −3.87, *P* = 0.002; bipolar peri-lesional target: t_11_ = −3.09, *P* = 0.010; focal peri-lesional target: t_11_ = −5.08, *P* < 0.001; Supp. Fig. 6B). Adding lesions also changed the 75% focality of |E| by a maximum of 50% (mean = 9.9%, SD = 13%), 9.0% (mean = 3.9%, SD = 3.1%), 30% (mean = 9.6%, SD = 8.9%), and 18% (mean = 5.8%, SD = 5.2%) for the bipolar M1, focal M1, bipolar peri-lesional target, and focal peri-lesional target, respectively (Supp. Fig. 7C). The focality metrics were both increased and decreased by adding lesions depending on participants (Supp. Fig. 7B).

#### Local metrics of Δmean |E|

Figure 3A shows the mean |E| in the ROIs for the healthy head models for reference. On average, tDCS induced a mean |E| of ∼0.2 V/m in the ROIs (bipolar M1: mean = 0.22 V/m, SD = 0.04 V/m; focal M1: mean = 0.20 V/m, SD = 0.08 V/m; bipolar peri-lesional target: mean = 0.20 V/m, SD = 0.04 V/m; focal peri-lesional target: mean = 0.19 V/m, SD = 0.07 V/m). The mean |E| was significantly higher in the bipolar M1 than focal M1 montage (t_12_ = 2.45, *P* = 0.031), but it was not significantly different between the bipolar and focal peri-lesional target montages.

**Figure 3.**
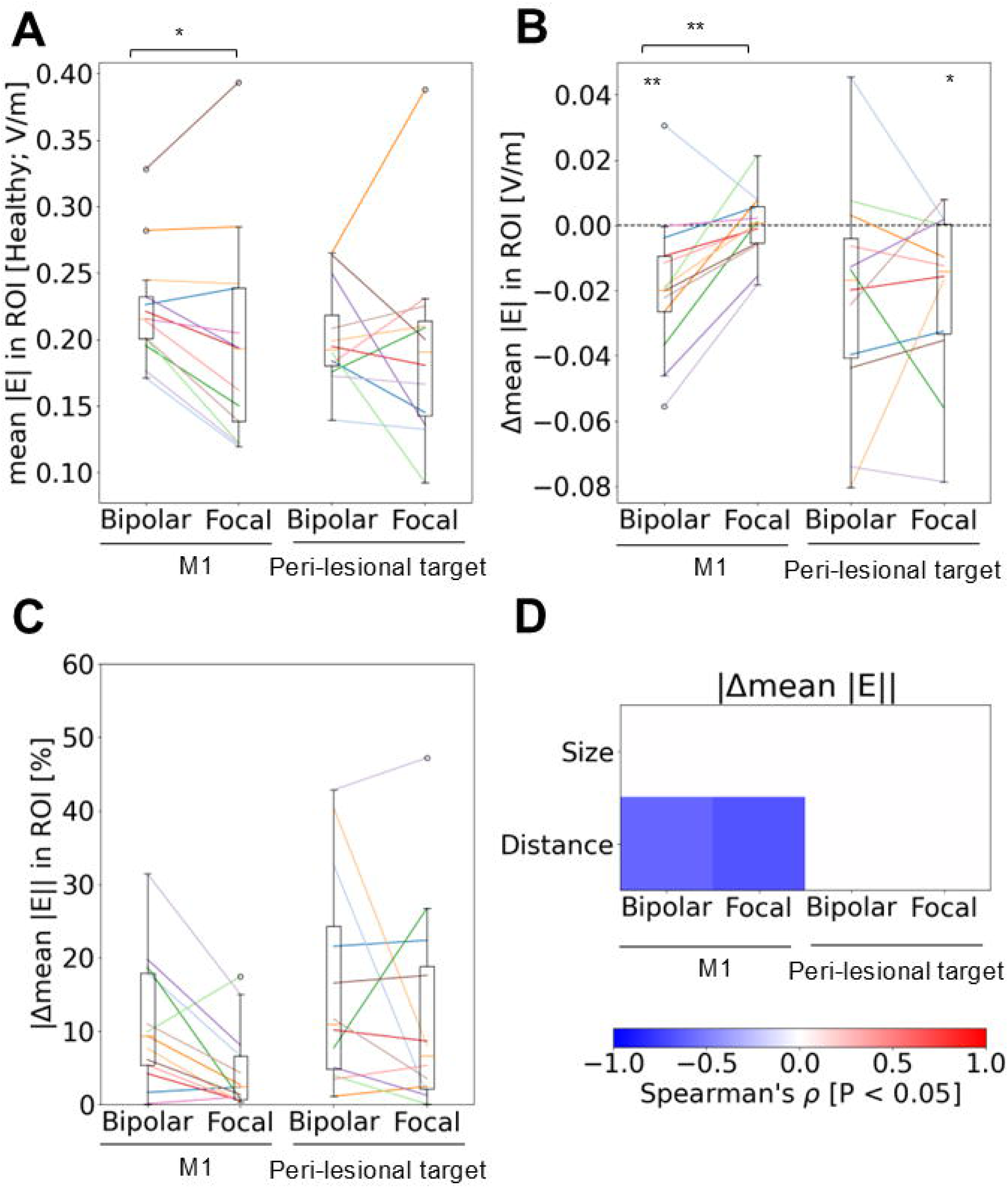
Change in the mean magnitude of electric field (mean |E|) within the region-of-interest (ROI) after adding artificial lesions to the healthy head models. (A-C) Boxplots of the mean |E| in ROI before adding artificial lesions (A) and the change after adding artificial lesions (B and C) for each montage (target location [the hand representation of left primary motor cortex (M1) or peri-lesional target] x type [bipolar or focal]). A and B show the actual value (V/m), while C indicates the relative absolute difference (%). (D) Spearman’s correlation coefficients between the absolute of Δmean |E| and the anatomical lesion features (the lesion size and lesion-to-target distance). Significant correlations (*P <* 0.05) are shown. * *P* < 0.05 ** *P* < 0.01

In general, adding lesions led to both decreases and increases in the mean |E| of the ROI (Fig. 3B) depending on the relative location of lesions (Fig. 4), which is in line with Evans et al., (2023). More specifically, adding lesions consistently decreased the mean |E| across participants in the bipolar M1 (mean = −0.018 V/m, SD = 0.021 V/m, t_12_ = 3.07, *P* = 0.010) and in the focal peri-lesional target (mean = −0.022 V/m, SD = 0.025 V/m, t_11_ = 2.72, *P* = 0.020) montages, while it was not consistently increased or decreased in the other montages. The differences corresponded to relative changes in the mean |E| of the ROI by a maximum 31% (mean = 11%, SD = 8.4%), 17% (mean = 4.7%, SD = 5.5%), 43% (mean = 16%, SD = 14%), and 47% (mean = 12%, SD = 14%) for the bipolar M1, focal M1, bipolar peri-lesional target, and focal peri-lesional target montages, respectively (Fig. 3C). The findings for nE were qualitatively similar to those reported here for |E| and are therefore reported in the *Supplementary information*.

**Figure 4.**
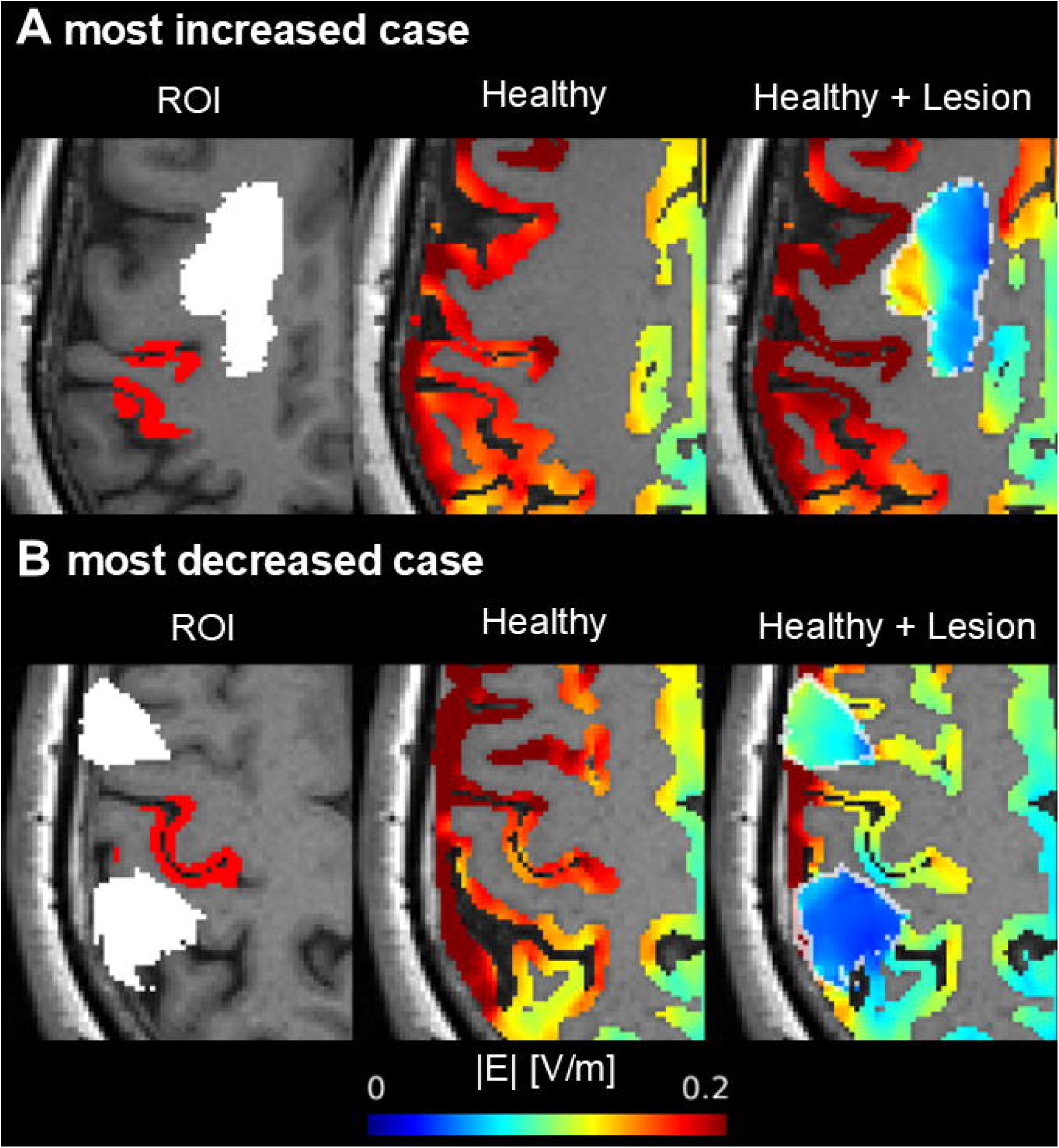
Simulated electric fields (E-fields) in representative healthy controls with bipolar montage targeting the left hand representation of primary motor cortex. (A) and (B) show the most increased and decreased, respectively, cases in mean magnitude of E-fields within the region-of-interest (ROI) after adding artificial lesions. The left panels show the location of ROI (red area) and lesions (white area). The middle panels show the simulated E-fields in the healthy head models, while the right panels represent those in the healthy head models with artificial lesions. The added lesions are surrounded by white lines.

We then explored associations between the differences in the mean |E| and the anatomical lesion features (Fig. 3D). The absolute change in the mean |E| due to the added lesions was significantly correlated to the lesion-to-target distance for both the bipolar and focal M1 montages (bipolar M1: r = −0.60, *P* = 0.031; focal M1: r = −0.67, *P* = 0.012). The other correlations were not significant.

#### Local metrics in mean |Δ|E||

The point-wise mean absolute difference in |E| within the ROI was significantly higher in the bipolar montage than the focal montage targeting M1 (t_12_ = 5.42, *P* < 0.001), while it was not significantly different across the montages targeting the peri-lesional target (Fig. 5A). Stated in percent of the baseline E-field without lesions, the mean |Δ|E|| within the ROI was up to 32% (mean = 13%, SD = 8.4%), 21% (mean = 7.5%, SD = 5.6%), 44% (mean = 23%, SD = 12%), and 47% (mean = 18%, SD = 11%) for the bipolar M1, focal M1, bipolar peri-lesional target, and focal peri-lesional target montages, respectively (Fig. 5B).

**Figure 5.**
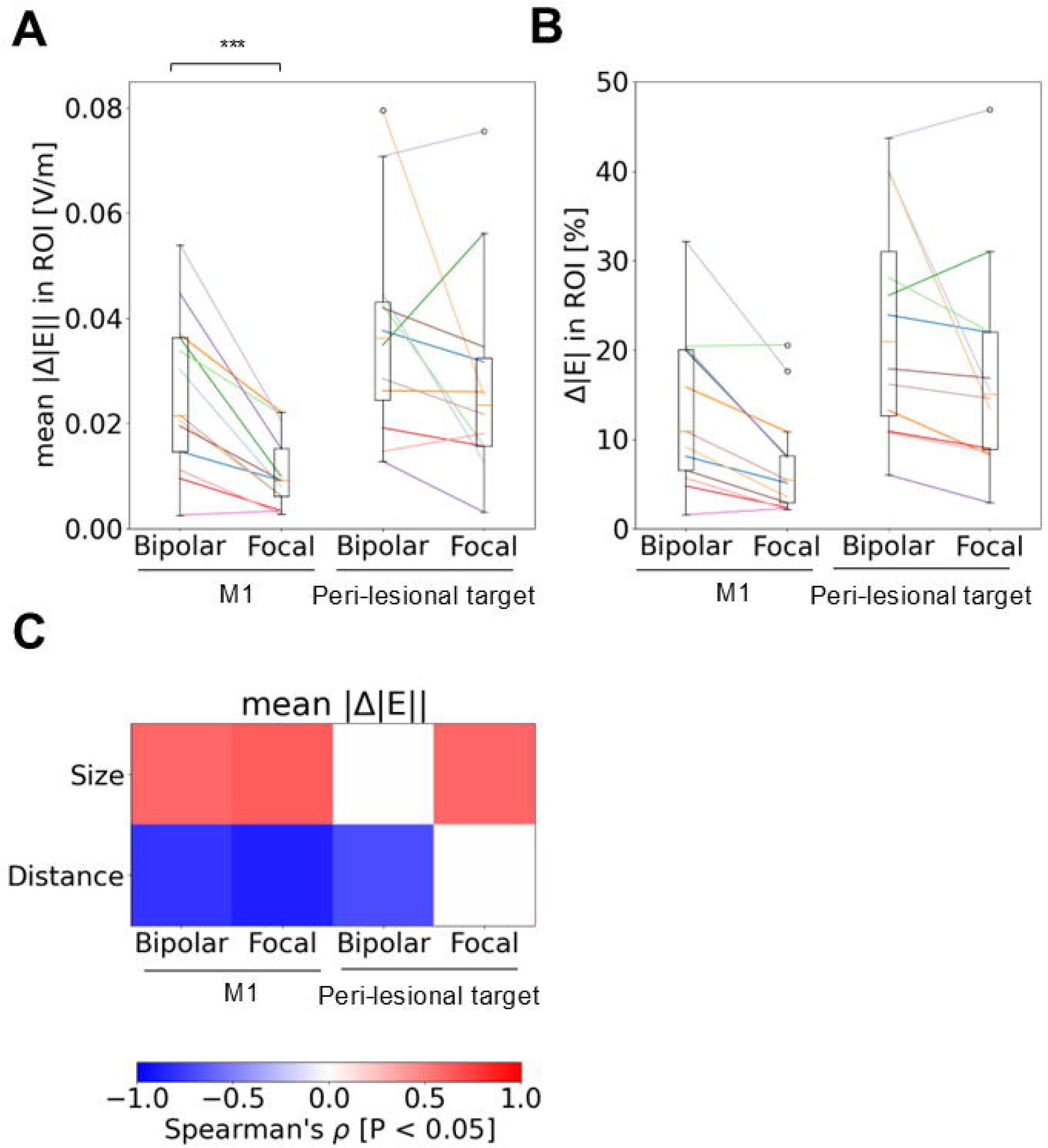
Position-wise comparisons of the magnitude of electric field (|E|) within the region-of-interest (ROI) after adding artificial lesions to the healthy head models. (A and B) Boxplots of the mean absolute difference in |E| (V/m, mean |Δ|E||, A) and the relative difference in |E| (%, B) within the ROI for each montage (target location [the hand representation of left primary motor cortex (M1) or peri-lesional target] x type [bipolar or focal]). (C) Spearman’s correlation coefficients between the mean |Δ|E|| and the anatomical lesion features (the lesion size and lesion-to-target distance). Significant correlations (*P <* 0.05) are shown. *** *P* < 0.001

Spearman’s correlation coefficients (Fig. 5C) demonstrated that the mean absolute difference in |E| was significantly positively correlated to the lesion size in three out of four montages (bipolar M1, r = 0.59, *P* = 0.032, focal M1, r = 0.64, *P* = 0.019, focal peri-lesional target, r = 0.59, *P* = 0.042). It was also significantly negatively correlated with the lesion-to-target distance in three montages (bipolar M1, r = −0.80, *P* = 0.001; focal M1, r = −0.87, *P* < 0.001; bipolar peri-lesional target: r = −0.70, *P* = 0.011). The remaining correlations were not significant.

### 3.2. Differences in E-fields between homogeneous- and realistic-lesion head models

The linear fits of conductivity against MD revealed consistent slope and intercept estimates across patients (slope: mean = 0.76 S•s/mm^3^, SD = 0.032 S•s/mm^3^, 0.68-0.80 S•s/mm^3^; intercept: mean = −0.40 S/m, SD = 0.061 S/m, −0.55 - −0.31 S/m; R^2^ [including WM and cCSF]: mean = 0.48, SD = 0.05, 0.37-0.55; see Supp. Fig. 8 for linear fits for a representative patient). Inside the lesion masks, the median lesion conductivities estimated via the diffusion-conductivity mapping for each patient showed substantial variability within and across patients (mean = 0.99 S/m, SD = 0.26 S/m, 0.48-1.38 S/m; Supp. Fig. 9). The lesion conductivities of the realistic-lesion head models were lower on average by 0.53-1.07 S/m (mean = 0.80 S/m, SD = 0.16 S/m).

#### Global metrics

We observed only small differences in the global metrics of the E-fields between the homogeneous- and realistic-lesion head models (peak 95% |E|: max = 10%, Supp Fig. 10; 75% focality |E|: max = 9.1%; Supp. Fig. 11).

#### Local metrics in mean |E|

We found small to moderate differences in the mean |E| within the ROI when changing from homogenous to realistic lesion modeling. The mean |E| in the baseline homogeneous lesion model was significantly higher in the bipolar than focal montages for both M1 (t_12_ = 4.67, *P* < 0.001) and peri-lesional targets (t_11_ = 3.18, *P* = 0.009; Fig. 6A). Modifying the lesion model resulted in a maximum of 12% (mean = 3.5%, SD = 3.6%), 4.4% (mean = 1.7%, SD = 1.5%), 23% (mean = 6.5%, SD = 5.7%), and 41% (mean = 6.3%, SD = 11%) difference in mean |E| within the ROI for the bipolar M1, focal M1, bipolar peri-lesional target, and focal peri-lesional target montages, respectively (Fig. 6C). The mean |E| within the ROI was significantly increased in the bipolar M1 montage (mean = 0.004 V/m, SD = 0.004 V/m, t_12_ = 3.45, *P* = 0.005), while no significantly consistent increases or decreases were observed in the other montages (Fig. 6B; see Figs. 7A and B for most increased and decreased case in |E| of the bipolar peri-lesional target montage, respectively). The decrease was significantly stronger in the bipolar M1 compared to the focal one (t_12_ = 5.74, *P* < 0.001), while it was not significantly different for the peri-lesional target (Fig. 6B).

**Figure 6.**
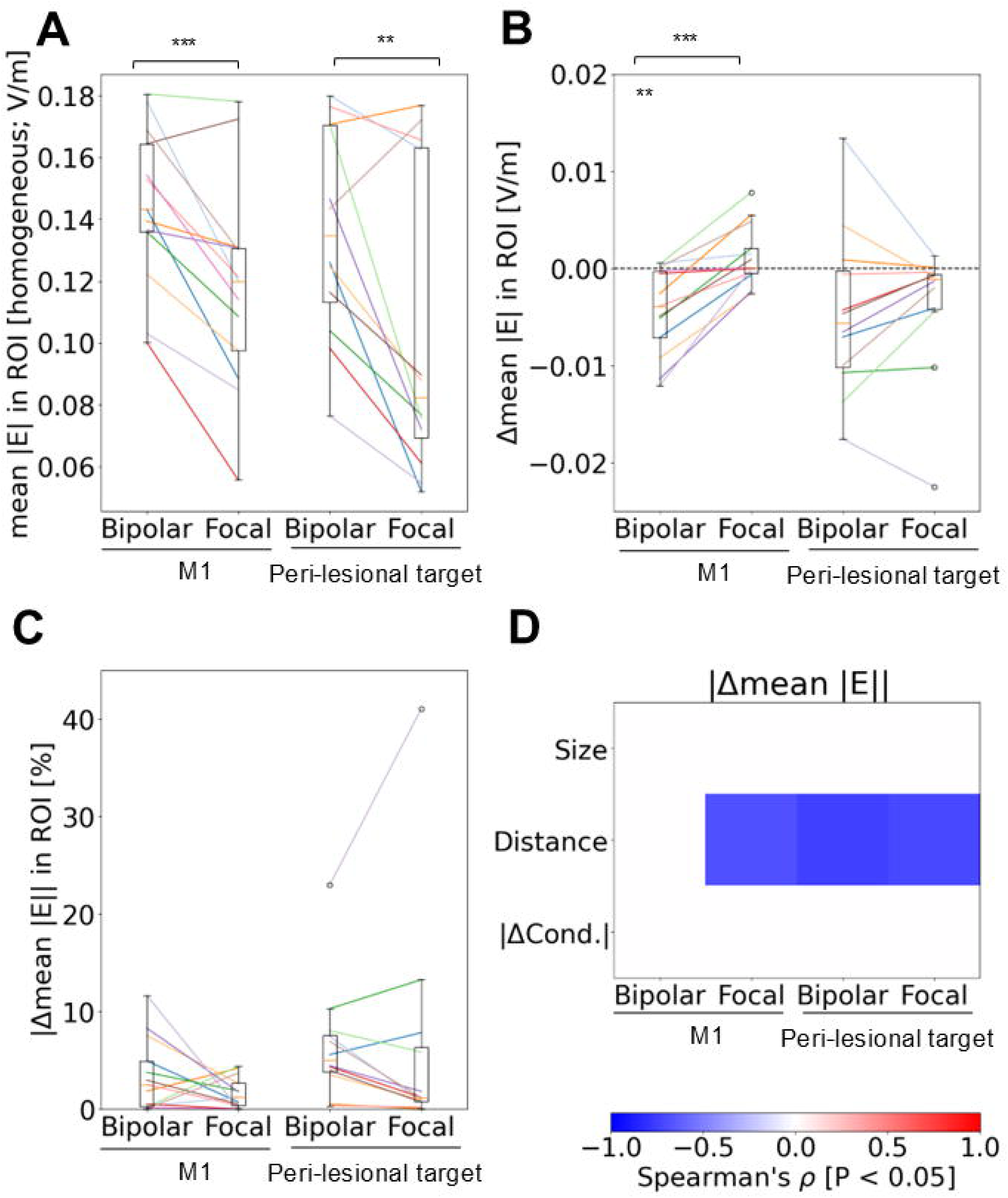
Change in the mean magnitude of electric field (mean |E|) within the region-of-interest (ROI) after changing the lesion conductivity from the homogeneous to realistic one in stroke patients. (A-C) Boxplots of the mean |E| in the ROI in the homogeneous-lesion head models (A) and the change after changing that head model to the realistic-lesion head models (B and C) for each montage (target location [the hand representation of left primary motor cortex (M1) or peri-lesional target] x type [bipolar or focal]). A and B show the actual value (V/m), while C indicates the relative absolute difference (%). (D) Spearman’s correlation coefficients between the absolute of Δmean |E| and the anatomical lesion features (the lesion size, lesion-to-target distance, and mean absolute difference of the conductivity within lesions [ΔCond.]). Significant correlations (*P <* 0.05) are shown. ** *P* < 0.01 *** *P* < 0.001

**Figure 7.**
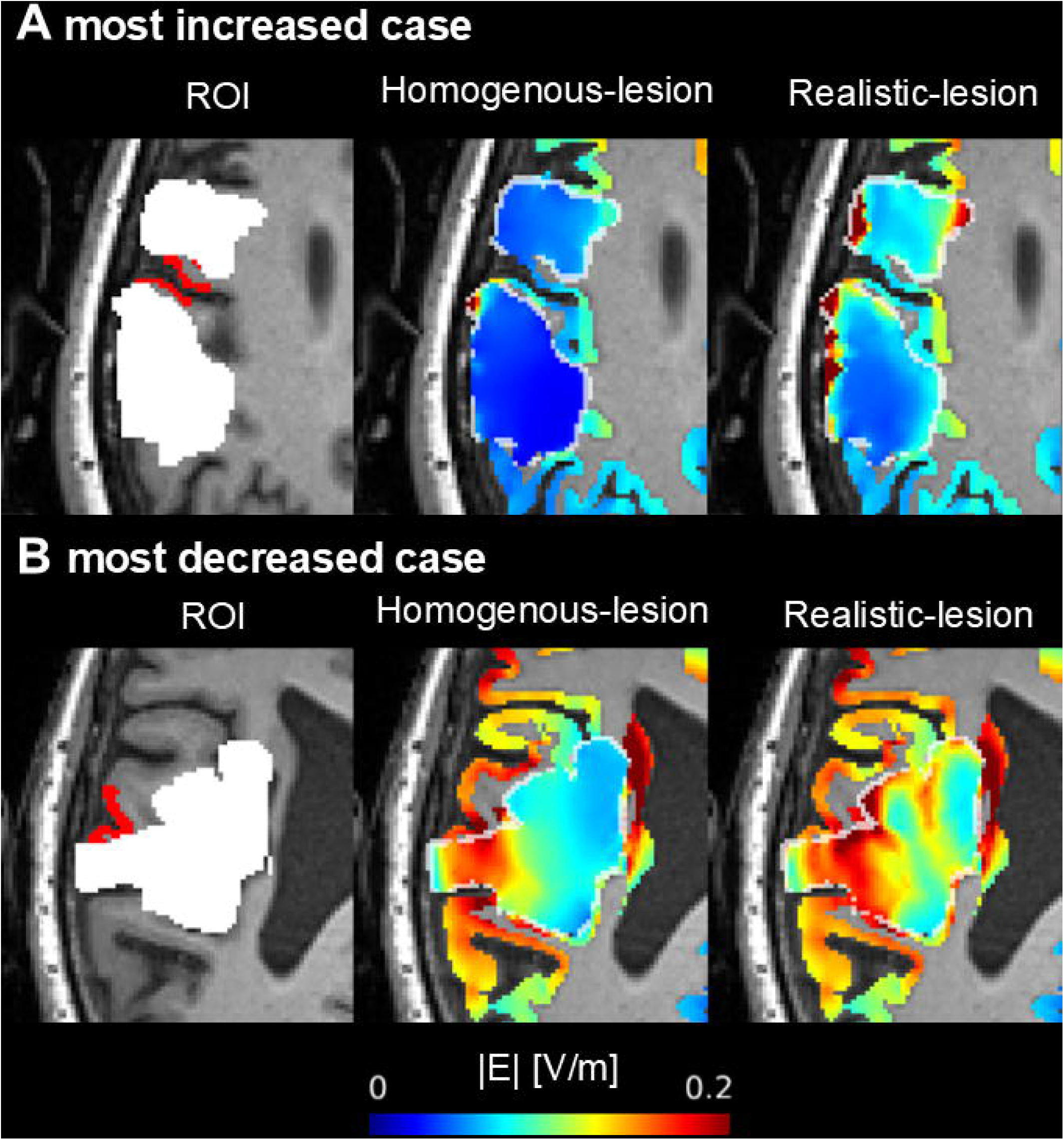
Simulated electric fields (E-fields) in representative stroke patients with bipolar montage targeting the peri-lesional target. (A) and (B) show the cases in which the mean magnitude of the E-fields within the region-of-interest (ROI) increased and decreased the most, respectively, after changing the lesion models from homogeneous to realistic. The left panels show the location of ROI (red area) and lesions (white area). The middle panels show the simulated E-fields in the homogeneous-lesion head models, while the right panels represent those in the realistic-lesion head models. The added lesions are surrounded by white lines.

Figure 6D illustrates the associations between the inter-individual variability of the difference in the mean |E| within the ROI and the anatomical lesion features. Here, we additionally investigated the correlation with the mean absolute difference in the conductivity within lesions because this value was highly variable across patients. The absolute difference in mean |E| between the two lesion models was significantly negatively correlated with the lesion-to-target distance in most montages (focal M1: r = −0.68, *P* = 0.010; bipolar peri-lesional target: r = −0.75, *P* = 0.005; focal peri-lesional target: r = −0.71, *P* = 0.009). In contrast, it was not significantly correlated to the lesion size or the mean absolute difference in the conductivity within lesions in any montage.

#### Local metrics in mean |Δ|E||

The mean absolute point-wise differences in |E| were small to moderate depending on individuals and montages (Fig. 8B). In percent, they were a maximum of 13% (mean = 5.8%, SD = 3.8%), 9.0% (mean = 3.4%, SD = 3.0%), 26% (mean = 9.8%, SD = 7.0%), and 40% (mean = 8.8%, SD = 11%) for the bipolar M1, focal M1, bipolar peri-lesional target, and focal peri-lesional target montages, respectively (Fig. 8B). The absolute differences in |E| were significantly higher in the bipolar than the focal montage for both the M1 (t_12_ = 5.15, *P* < 0.001) and peri-lesional targets (t_11_ = 3.09, *P* = 0.010; Fig. 8A).

**Figure 8.**
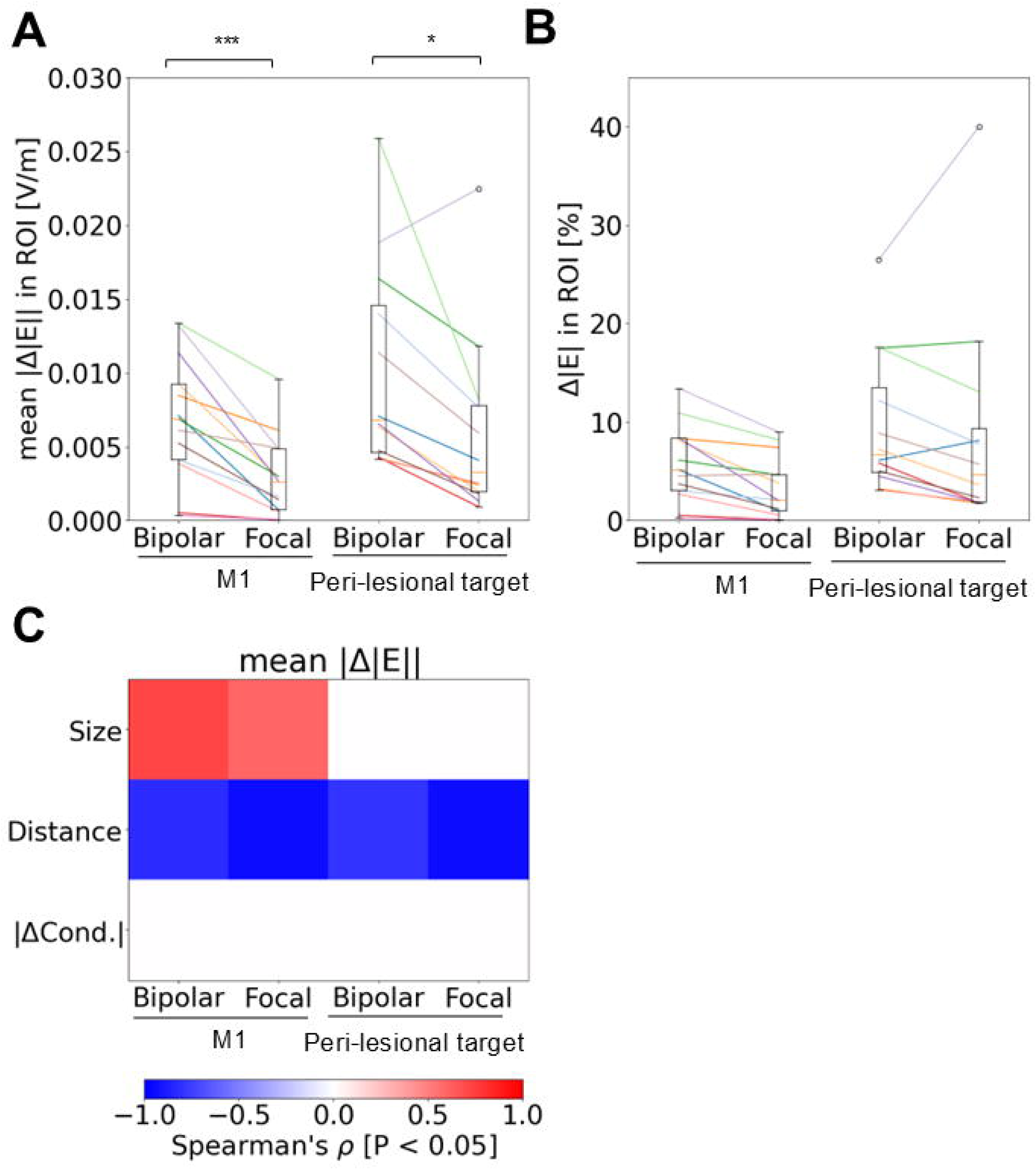
Position-wise comparisons of the magnitude of electric field (|E|) within the region-of-interest (ROI) after changing the lesion conductivity from homogeneous to realistic in stroke patients. (A and B) Boxplots of the mean absolute difference in |E| (V/m, mean |Δ|E||, A) and the relative difference in |E| (%, B) within the ROI for each montage (target location [the hand representation of left primary motor cortex (M1) or peri-lesional target] x type [bipolar or focal]). (C) Spearman’s correlation coefficients between the mean |Δ|E|| and the anatomical lesion features (the lesion size, lesion-to-target distance, and mean absolute difference of the conductivity within the lesions [ΔCond.]). Significant correlations (*P <* 0.05) are shown. * *P* < 0.05, *** *P* < 0.001

Figure 8C shows the correlations between the inter-individual variability of the point-wise mean absolute difference in |E| in the ROI and the anatomical lesion features. The mean |Δ|*E||* was significantly positively correlated to the lesion size in the two M1 montages (bipolar M1: r = 0.72, *P* = 0.006; focal M1: r = 0.60, *P* = 0.031). It was significantly negatively correlated to the lesion-to-target distance in all montages (bipolar M1: r = −0.82, *P* < 0.001; focal M1: r = −0.94, *P* < 0.001; bipolar peri-lesional target: r = −0.79, *P* = 0.002; focal peri-lesional target: r = −0.94, *P* < 0.001).

## 4. Discussion

In this study, we evaluated the impact of lesions on tDCS-induced E-fields using realistic size and shape of lesion masks from stroke patients. We also assessed the extent to which modeling spatial heterogeneity of lesion conductivity impacts the simulated E-field. Compared to prior studies (Datta et al., 2011; Evans et al., 2023; Galletta et al., 2015; Krishnamurthy et al., 2025; Richardson et al., 2015; Minjoli et al., 2017), the use of a larger dataset that also included diffusion MRI enabled us to relate interindividual differences in lesion size and composition to the resulting effects on the tDCS-induced E-fields. We found that adding lesions to a healthy head model can affect the |E| near the targeted region by up to 47% depending on the size of the lesions and the distance of a lesion from the target location. Diffusion-to-conductivity mapping (realistic-lesion head model) revealed high variability in lesion conductivity within and across the stroke patients, and incorporating heterogeneous conductivity introduced additional small to moderate differences in |E| nearby the target location compared to homogeneous-lesion head models. These differences were most pronounced for bipolar montages, larger lesional volumes, and closer locations of the lesions relative to the stimulation target. Considering that lesions in patients after stroke are often close to the target area, as in our dataset on stroke-induced aphasia (e.g., Fig. 4B), we recommend acquiring dMRI data prior to tDCS to apply realistic-lesion head models in future stroke studies employing prospective or retrospective modeling of tDCS-induced E-fields.

### 4.1. Lesions can both increase and decrease |E| nearby the target region

Adding homogenous lesions to a heathy head model led to both increases and decreases in |E| within the ROI. This aligns with a previous simulation study that systematically placed artificial lesions around the ROI. That study showed that |E| was increased when the lesion was aligned with the main E-field direction and decreased when located in its orthogonal direction (Evans et al., 2023). Our results extended this previous finding by showing that such increase or decrease can also occur for realistic lesion shapes and lesion-to-target distances in stroke patients. These findings indicate that the direction of lesion impact on |E| cannot be predicted without simulation and underscore the need for using individual head models to individually optimize tDCS parameters in stroke patients.

Minjoli et al., (2017) reported a reduction of the average cortical field strength in two stroke patients compared to a healthy control. While this is in line with the reduction of the 95% peak |E| that was observed here in most cases when adding an artificial lesion to the healthy controls, we observed that |E| can both increase and decrease within the ROI. We speculate that further anatomical differences between the brains of the healthy control and stroke patients might have additionally contributed to the marked difference seen in Minjoli et al., (2017). In particular, increased atrophy levels in the stroke patients might have promoted current shunting through CSF, further reducing the E-field in gray matter.

Adding homogeneous lesions altered |E| within the ROI by an average of 7.5-23% depending on the electrode montage. This change is large given that a previous simulation study reported an average change in |E| of ∼5% within the ROI when artificial lesions were added to healthy head models (Evans et al., 2023). This difference is probably due to the difference in the investigated lesion size and the lesion-to-target distance. In Evans et al., (2023), artificial lesions were added with spheres ranging from 4 to 24 mm in radius (268-57,906 mm^3^), while the lesional volume in our study was larger (7,181-110,304 mm^3^). Furthermore, while that study did not consider the case that lesions overlap with the ROI, we found that this can happen in both M1 and peri-lesional target (lesion-to-target distance < 12.5 mm in Suppl. Fig. 5B). Considering that the mean pointwise absolute difference in |E| within the ROI was positively and negatively correlated to the lesion size and lesion-to-target distance, respectively, in most montages, these factors would define the degree of the effect of lesions in tDCS-induced E-field. In summary, we found that, even with the assumption of homogeneous lesions, these lesions can moderately increase or decrease the E-field, highlighting the need for personalized head models for stroke patients.

### 4.2. Diffusion-to-conductivity mapping revealed substantial heterogeneity in the conductivity of lesions across the patients

One further important aspect is the heterogeneity of the lesion conductivity within and across the stroke patients. Our diffusion-to-conductivity mapping method indeed revealed high intra- and inter-individual variability in the lesion conductivities. A previous meta-analysis on lesion conductivity also showed large variability in the lesion conductivity across five studies (0.10-1.77 S/m; McCann et al., (2019)). Together with our results, these observations suggest that lesion conductivity values should be individualized based on lesion properties to improve the simulation accuracy of E-field in stroke patients. The meta-analysis also reported that the weighted mean of the conductivity of lesions across the studies was 0.88 S/m [SD = 0.38 S/m]. This value is comparable to the group-averaged median conductivity within the lesions in our study (0.99 S/m [SD = 0.26 S/m]), supporting the validity of our method.

### 4.3. The degree of the difference in |E| between homogeneous- and realistic-lesion model was dependent on the lesion size and lesion-to-target distance

Mean absolute |E| differences were positively correlated with lesion size in several montages. This is consistent with a previous simulation study (Evans et al., 2023), and expected because the differences in the lesion conductivities between the two head models are more widespread if the lesions are large. The more widespread differences in conductivity would make the E-field within the ROI more susceptible to the differences in lesion model. Thus, using realistic lesion modeling can be more important as the lesion size increases.

In addition, these differences were negatively correlated with the lesion-to-target distance across all montages. As in previous (Evans et al., 2023) and our studies, the effect of lesions on the simulated E-field within the ROI is larger if the lesions are closer to that ROI. Therefore, the E-field within the ROI would be also more affected by the differences in lesion model with shorter lesion-to-target distance. Given that significant correlations were observed in lesion-to-target distance more consistently than in lesion size, the lesion proximity may be more critical determinant of the |E| differences between the homogenous- and realistic-lesion head models than lesion size.

### 4.4. Modifying the lesion model had a larger impact on |E| more in the bipolar montages than focal montages

The mean absolute change in |E| within the ROI after modifying the lesion model from homogeneous to realistic was consistently larger in the bipolar than in focal montages. This likely reflects the fact that bipolar montages distribute tDCS-induced current more broadly within the brain than focal montages. Consequently, lesions located anywhere nearby this more widespread current pathway in bipolar montage would be more likely to influence current flow than focal montages. These findings suggest that montage selection is another key determinant of whether realistic-lesion modeling substantially alters simulated E-fields, and that bipolar montages, in particular, warrant cautious head modeling in stroke populations.

### 4.5. Future directions

Although the realistic-lesion head model employed in this study is promising for individual simulations of tDCS-induced current in stroke patients, the underlying diffusion-to-conductivity mapping has not yet been validated. This could be achieved by measurements, e.g. based on magnetic resonance current density imaging (Gregersen et al., 2024), to support the accuracy of the estimated E-field from the realistic-lesion head model.

Beyond methodological validation, establishing the clinical utility of our approach will be relevant as well. Specifically, it remains to be demonstrated whether and how replacing homogeneous-with realistic-lesion head models benefits neurorehabilitation studies in stroke patients. For instance, simulation of tDCS-induced E-fields can be used to probe the link between E-field and behavioral outcomes (Antonenko et al., 2019) or to optimize stimulation parameters of tDCS (Datta et al., 2011; Galletta et al., 2015). Future studies could therefore address two key questions: (1) whether E-fields derived from realistic-lesion head models show higher correlation to neurophysiological and behavioral outcomes in stroke patients than those from homogeneous-lesion head model; and (2) in prospective approach, whether optimizing stimulation parameters using realistic-lesion head models enhances rehabilitation outcomes more effectively and robustly than homogenous-lesion head models.

### 4.6. Conclusions

Our study demonstrated that homogenous-lesion head models can alter |E| nearby the target location by up to 47% compared to healthy head models. Our diffusion-to-conductivity mapping approach revealed substantial variability in the conductivity of lesions both within and across stroke patients. Modifying the lesion model from homogeneous to realistic produced small to moderate differences, particularly when bipolar montages were applied, lesions were larger, or lesions were located closer to the target region. These findings indicate that realistic-lesion head model should be employed to simulate tDCS-induced E-field and design individualized stimulation protocols accurately for stroke patients. Incorporating this model may enhance the precision and efficacy of personalized tDCS interventions in stroke rehabilitation, an approach to be validated in future prospective studies.

## Data Availability

The processed data of this study are available upon request from the corresponding author. The raw data are not publicly available due to potential identifying information that could compromise participant privacy.

## CRediT authorship contribution statement

**Ikko Kimura:** Methodology, Software, Validation, Formal analysis, Writing-Original draft preparation, Visualization; **Marcus Meinzer**: Investigation, Writing-Original draft preparation, Writing-Reviewing and Editing; **Daria Antonenko**: Methodology, Data Curation, Writing-Reviewing and Editing; **Robert Darkow**: Investigation, Data Curation, Writing-Reviewing and Editing; **Agnes Flöel**: Writing-Reviewing and Editing, Supervision, Project administration, Funding acquisition; **Axel Thielscher**: Conceptualization, Methodology, Writing-Reviewing and Editing, Supervision, Project administration, Funding acquisition

## Acknowledgments

This work was supported by the German Research Foundation (Research Unit 5429/1 (467143400), AF 379/34-1, AF 379/35-1, MM 3161/5-1, MM 3161/6-1, DA 1103/5-1, AT 1330/6-1, AT 1330/7-1), and Bundesministerium für Bildung und Forschung (AF: FKZ0315673A, 01GY1144; 01GQ1424A, 01GQ1420B; AF and MM: 01EO0801). DA was supported by the Heisenberg Programme of the German Research Foundation (project number: 539593253). AT was supported by the Lundbeck foundation (grant R313-2019-622).

## Declaration of generative AI and AI-assisted technologies in the writing process

During the preparation of this manuscript, the authors used language editing tools (ChatGPT and DeepL) to improve readability and clarity. All content was subsequently reviewed and revised by the authors, who take full responsibility for the final version of the article.

