## Supplementary Information for "Simulating tDCS-induced electric fields in stroke patients: realistic-lesion head models are needed"

**Title Page**

**Affiliations**
^1^ Danish Research Centre for Magnetic Resonance, Department of Radiology and Nuclear Medicine, Copenhagen University Hospital - Amager and Hvidovre, Copenhagen, Denmark
^2^ Department of Neurology, University Medicine Greifswald, Greifswald, Germany

^3^ FH Joanneum Gesellschaft mbH, Graz, Australia
^4^ German Centre for Neurodegenerative Diseases (DZNE) Standort Greifswald, Greifswald, Germany ^5^ Section for Magnetic Resonance, DTU Health Tech, Technical University of Denmark, Kgs Lyngby, Denmark

^†^ Equally contributed first authors

^‡^ Equally contributed last authors

Contains Supplementary Results and Supplementary Figures 1-11

**Supplementary Results**

Differences in nE between healthy head models with and without artificial lesions

*Local metrics in* $\Delta$*mean nE*: Adding homogeneous lesions to healthy head models decreased the mean nE within the ROI in most cases, while increases occurred in some participants. The mean nE of the healthy head models was not significantly different across the bipolar and focal montages for either M1 or peri-lesional target (Supp. Fig. 1A). The decreases were significant for the bipolar M1 montage (t_12_ = -2.28, *P* = 0.042), but not for the other montages (Supp. Fig. 1B). The decrease of the bipolar M1 montage was significantly larger than for the focal M1 montage (t_12_ = 2.81, *P* = 0.016). The relative differences in the mean nE in the ROI exhibited maximums of 35% (mean = 10%, SD = 8.9%), 5.7% (mean = 2.5%, SD = 2.1%), 46% (mean = 18%, SD = 14%), and 43% (mean = 9.1%, SD = 12%) for the bipolar M1, focal M1, bipolar peri-lesional target, and focal peri-lesional target montages, respectively (Supp. Fig. 1C).

*Local metrics in mean |*$\Delta$*nE|*: The mean absolute difference in nE was significantly higher for the bipolar than the focal montages for both the M1 (t_12_ = 4.82, *P* < 0.001) and peri-lesional targets (t_11_ = 2.94, *P* = 0.013; Supp. Fig. 2A). The relative difference in nE revealed up to 69% (mean = 27%, SD = 18%), 65% (mean = 16%, SD = 16%), 87% (mean = 40%, SD = 19%), and 69% (mean = 30%, SD = 16%) differences for the bipolar M1, focal M1, bipolar peri-lesional target, and focal peri-lesional target montages, respectively (Supp. Fig. 2B).

Differences in nE between homogeneous- and realistic-lesion head models

*Local metrics in* $\Delta$*mean nE*: For mean nE, we found maximum 9.8% (mean = 5.0%, SD = 3.3%), 4.1% (mean = 1.7%, SD = 1.3%), 20% (mean = 5.9%, SD = 4.9%), and 28% (mean = 4.4%, SD = 7.5%) difference between the homogeneous- and realistic-lesion head models for the bipolar M1, focal M1, bipolar peri-lesional target, and focal peri-lesional target montages, respectively (Supp. Fig. 3C). Changing the lesion model from the homogeneous to realistic significantly increased the mean nE within the ROI in the bipolar M1 montage (t_12_ = 5.41, *P* < 0.001), while there was no consistent increase or decrease in the other montages (Supp. Fig. 3B). The difference for the bipolar M1 montage was significantly higher than that for the focal M1 montage (t_12_ = 5.79, *P* < 0.001), while it was significantly different for the peri-lesional montages (Supp. Fig. 3B). The mean nE of the homogeneous-lesion head model was significantly higher for the bipolar montages than the focal ones for both the M1 (t_12_ = 6.10, *P* < 0.001) and peri-lesional targets (t_11_ = 3.52, *P* = 0.005; Supp. Fig. 3A).

*Local metrics in mean |*$\Delta$*nE|*: Stated in percent, we observed maximum differences in nE of 24% (mean = 10%, SD = 6.8%), 18% (mean = 7.4%, SD = 6.3%), 26% (mean = 14%, SD = 7.4%), and 34% (mean = 13%, SD = 9.2%) between the homogeneous- and realistic-lesion head models for the bipolar M1, focal M1, bipolar peri-lesional target, and focal peri-lesional target montages, respectively (Supp. Fig. 4A). The mean absolute differences of nE within the ROI were higher for the bipolar than the focal montages for both the M1 (t_12_ = 4.63, *P* < 0.001) and peri-lesional targets (t_11_ = 3.89, *P* = 0.003; Supp. Fig. 4B).

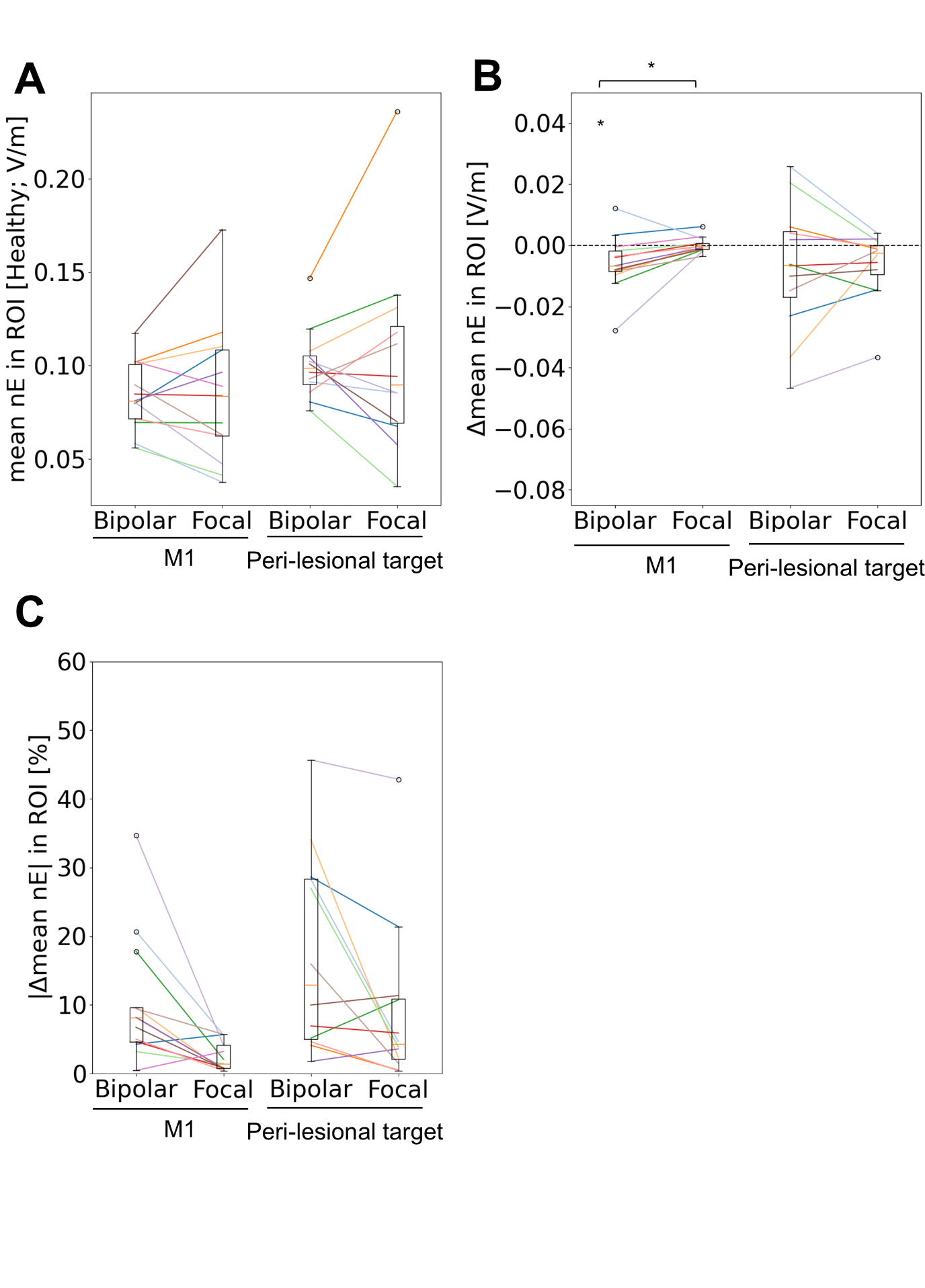
**Supplementary Figure 1. Changes in the mean normal component of E-fields (nE) within the region-of-interest (ROI) after adding artificial lesions to the healthy head models.** Boxplots of the mean nE in the healthy head models (A) and the difference in nE ($\Delta$mean nE; B and C) for each montage (target location [the hand representation of left primary motor cortex (M1) or peri-lesional target] x type [bipolar or focal]). A and B show the actual value (V/m), while C indicates the relative absolute difference (%). * *P* < 0.05

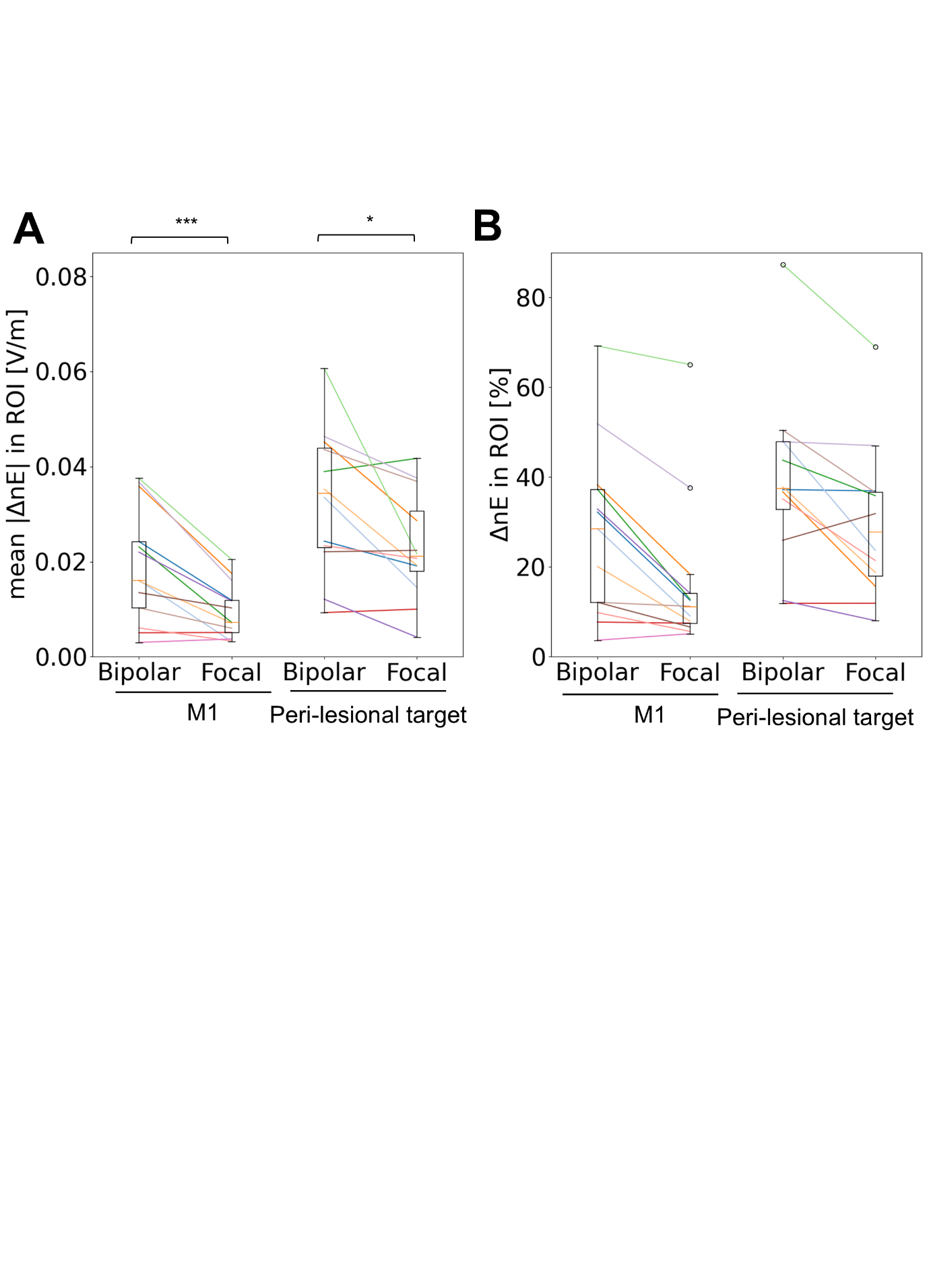

**Supplementary Figure 2. Position-wise comparisons of the normal component of electric field (nE) within the region-of-interest (ROI) after adding artificial lesions to the healthy head models.** (A and B) Boxplots of the mean absolute difference in nE (V/m, mean |$\Delta$nE|, A) and the relative difference in nE (%, B) within the ROI for each montage (target location [the hand representation of left primary motor cortex (M1) or peri-lesional target] x type [bipolar or focal]). * *P* < 0.05, *** *P* < 0.001

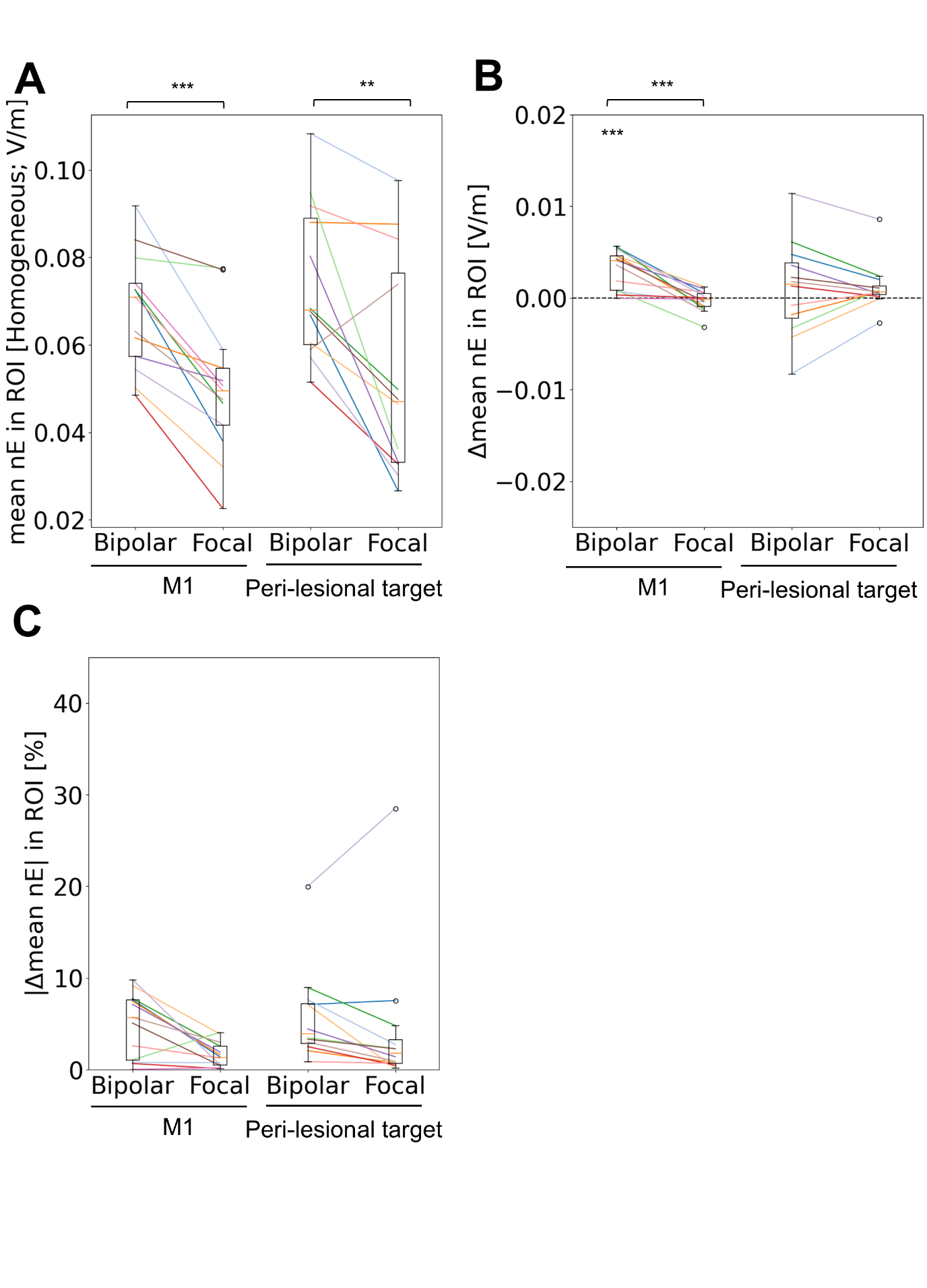

**Supplementary Figure 3. Change in the mean normal component of electric field (mean nE) within the region-of-interest (ROI) after changing the lesion conductivity from homogeneous to realistic in stroke patients.** (A-C) Boxplots of the mean nE in the ROI in the homogeneous-lesion head models (A) and the change after changing that head model to the realistic-lesion head models (B and C) for each montage (target location [the hand representation of left primary motor cortex (M1) or peri-lesional target] x type [bipolar or focal]). A and B show the actual value (V/m), while C indicates the relative absolute difference (%). ** *P* < 0.01 *** *P* < 0.001

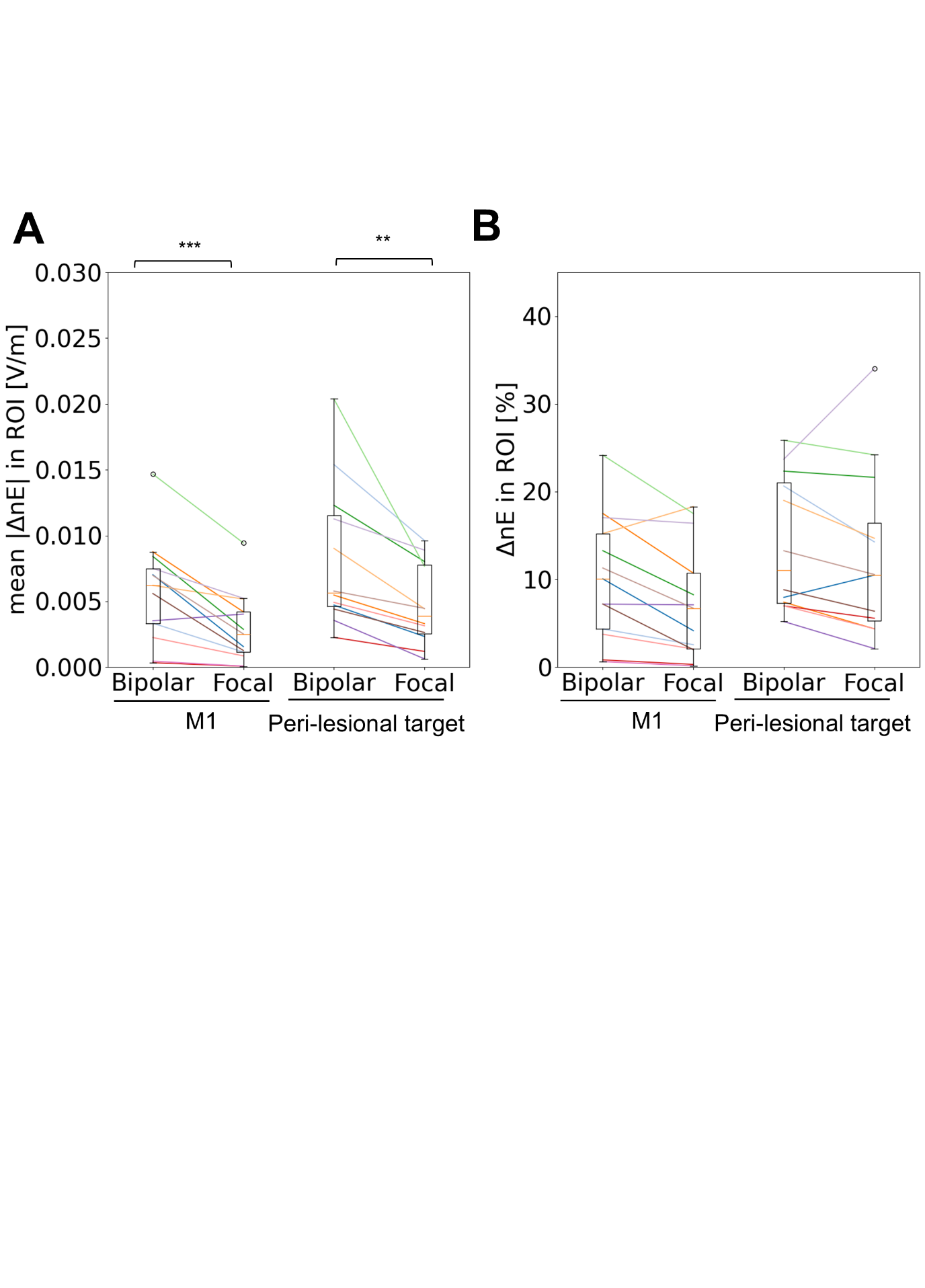

**Supplementary Figure 4. Position-wise comparisons of the normal component of electric field (nE) within the region-of-interest (ROI) after changing the lesion conductivity from homogeneous to realistic in stroke patients.** (A and B) Boxplots of the mean absolute difference in nE (V/m, mean |$\Delta$nE|, A) and the relative difference in nE (%, B) within the ROI for each montage (target location [the hand representation of left primary motor cortex (M1) or peri-lesional target] x type [bipolar or focal]). ** *P* < 0.01, *** *P* < 0.001

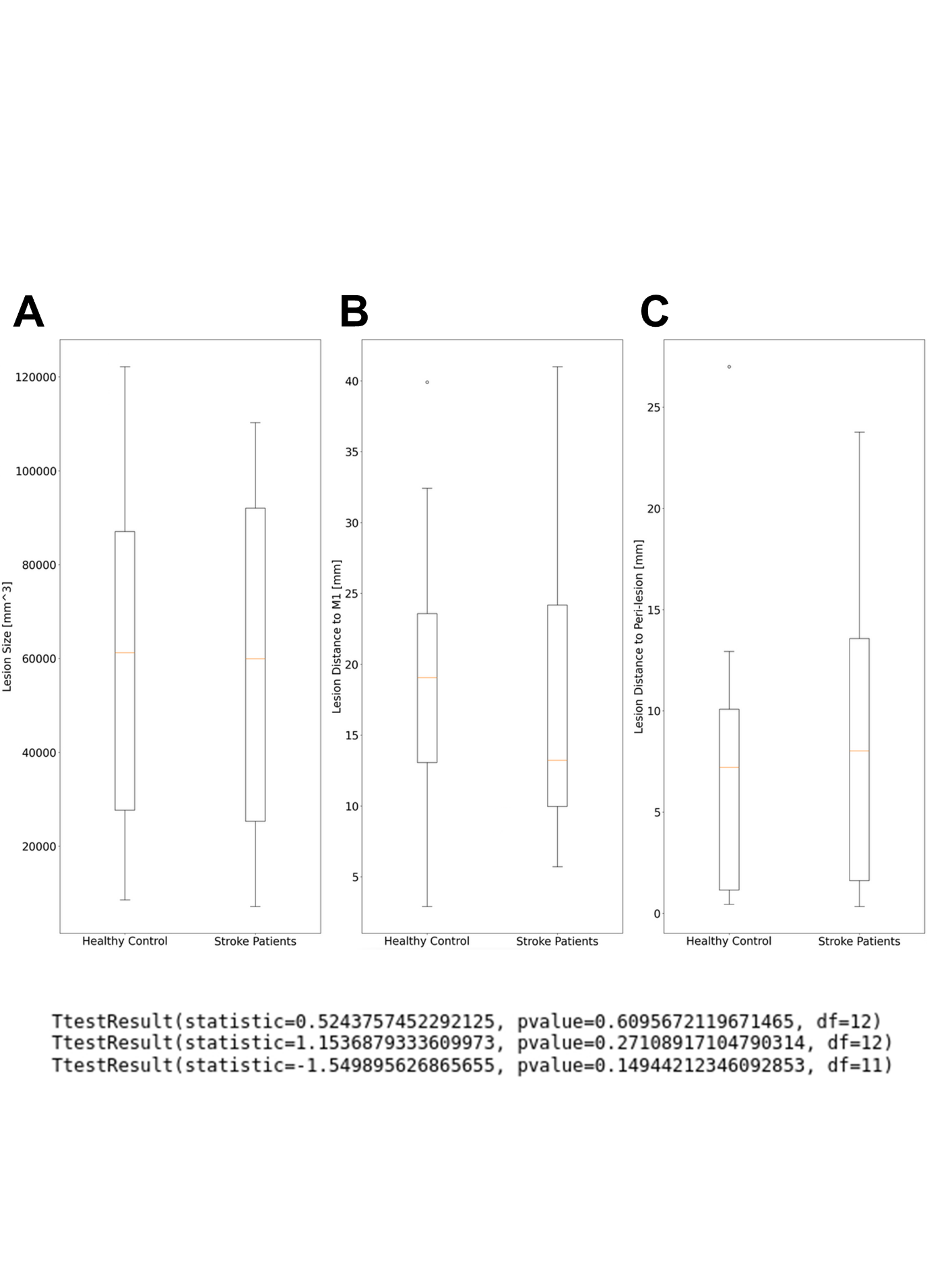

**Supplementary Figure 5. Anatomical lesion features tested across healthy controls and stroke patients.** Boxplots of lesion size (A), lesion distance to the hand representation of left primary motor cortex (M1; B), and lesion distance to the peri-lesional target (C). The artificial lesions tested in the healthy controls were non-linearly transformed from the lesions of the age-matched pairs of the stroke patients (See Section 2.4. in the main text for details).

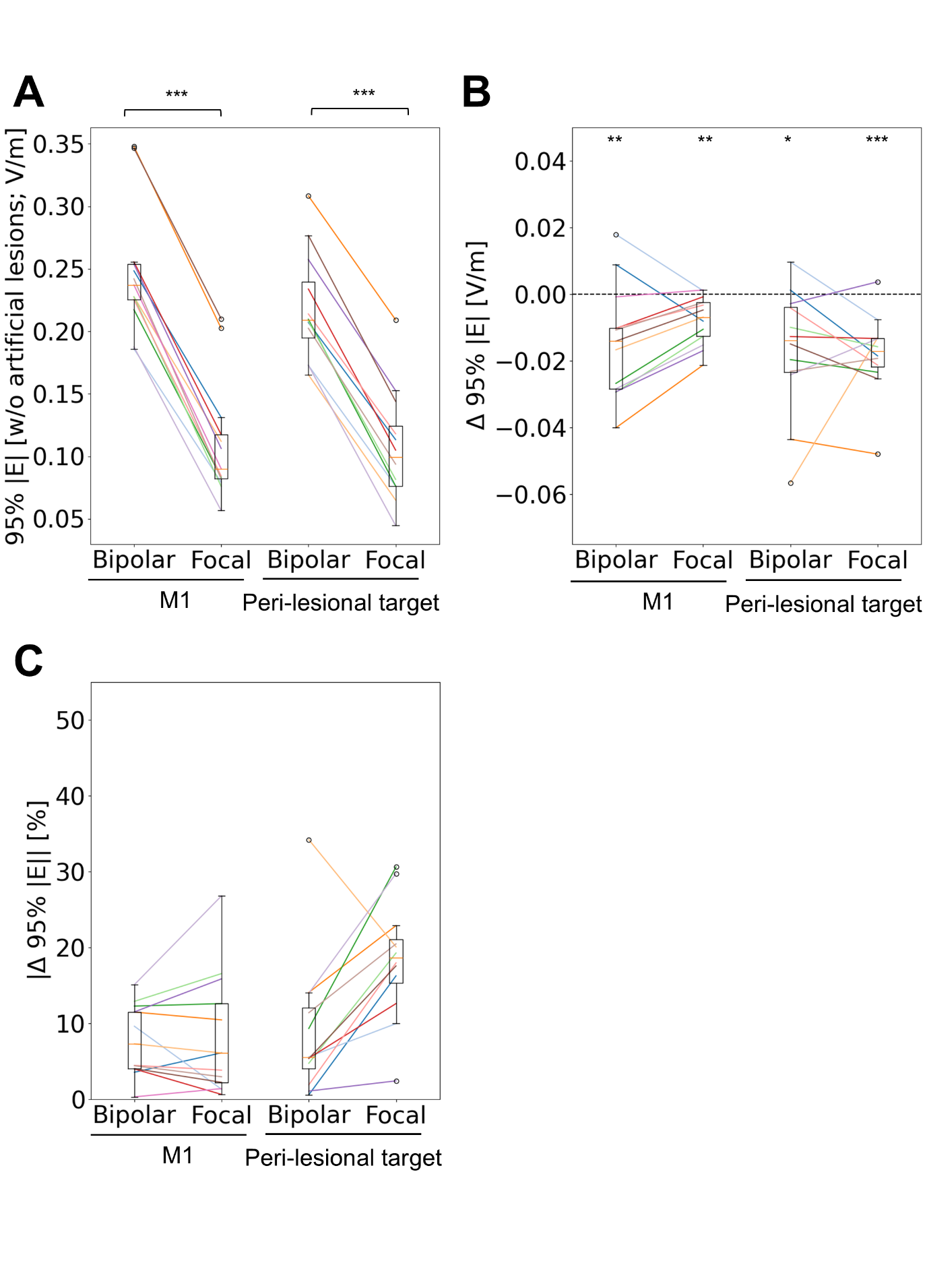

**Supplementary Figure 6. Change in the 95^th^ percentile peak magnitude of electric field (95% |E|) within the gray matter after adding artificial lesions to the healthy head models.** (A-C) Boxplots of 95% peak |E| in the healthy head models (A) and the change after adding artificial lesions (B and C) for each montage (target location [the hand representation of left primary motor cortex (M1) or peri-lesional target] x type [bipolar or focal]). A and B show the actual value (V/m), while C indicates the relative absolute difference (%). * *P* < 0.05 ** *P* < 0.01 *** *P* < 0.001

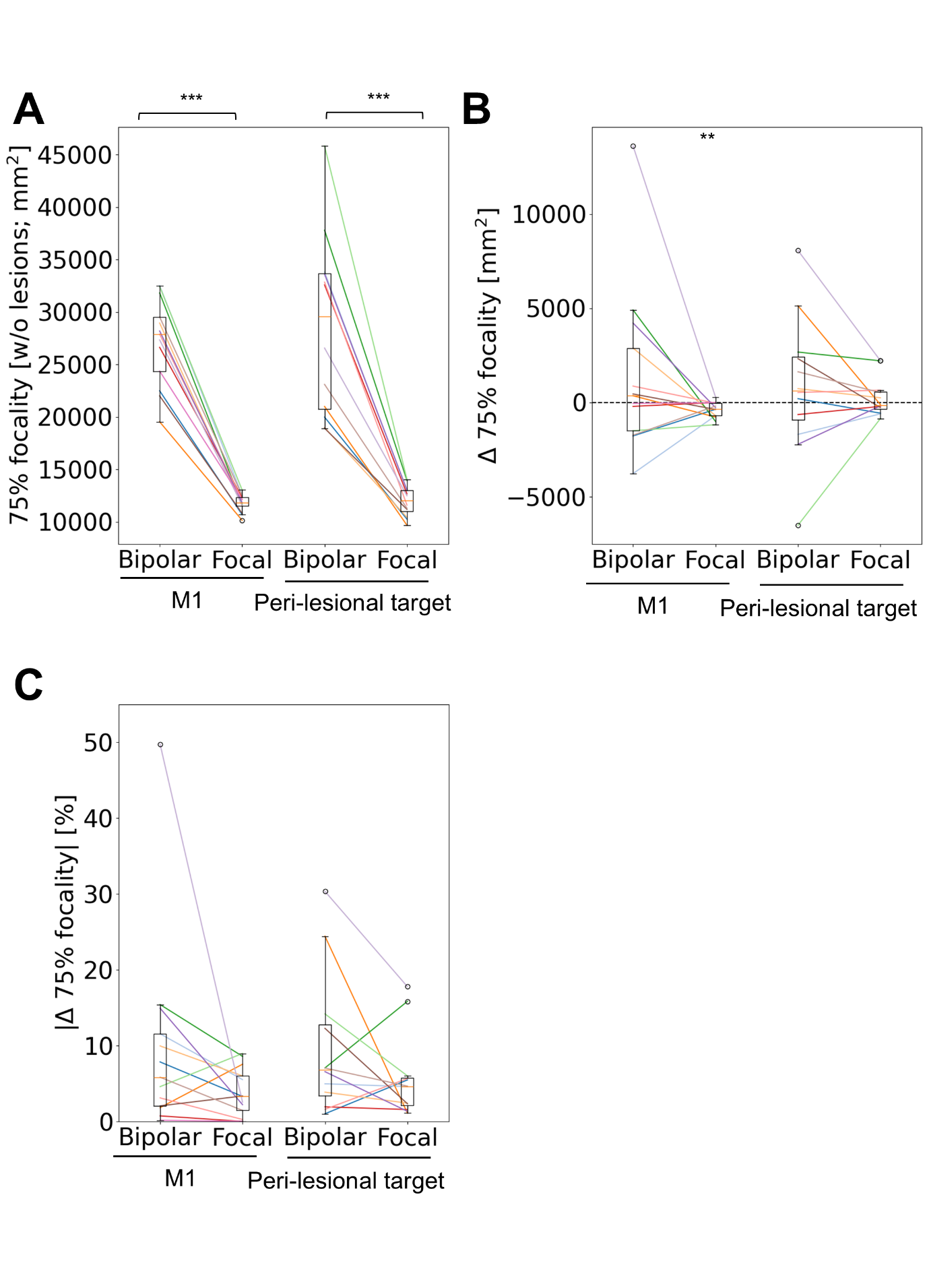

**Supplementary Figure 7. Change in the focality of electric field (E-field) within the gray matter after adding artificial lesions to the healthy head models.** Focality was defined as the area more than 75% above the 95^th^ percentile of the magnitude of E-field (75% focality). (A-C) Boxplots of 75% focality in the healthy head models (A) and the change after adding artificial lesions (B and C) for each montage (target location [the hand representation of left primary motor cortex (M1) or peri-lesional target] x type [bipolar or focal]). A and B show the actual value (mm^2^), while C indicates the relative absolute difference (%). ** *P* < 0.01 *** *P* < 0.001

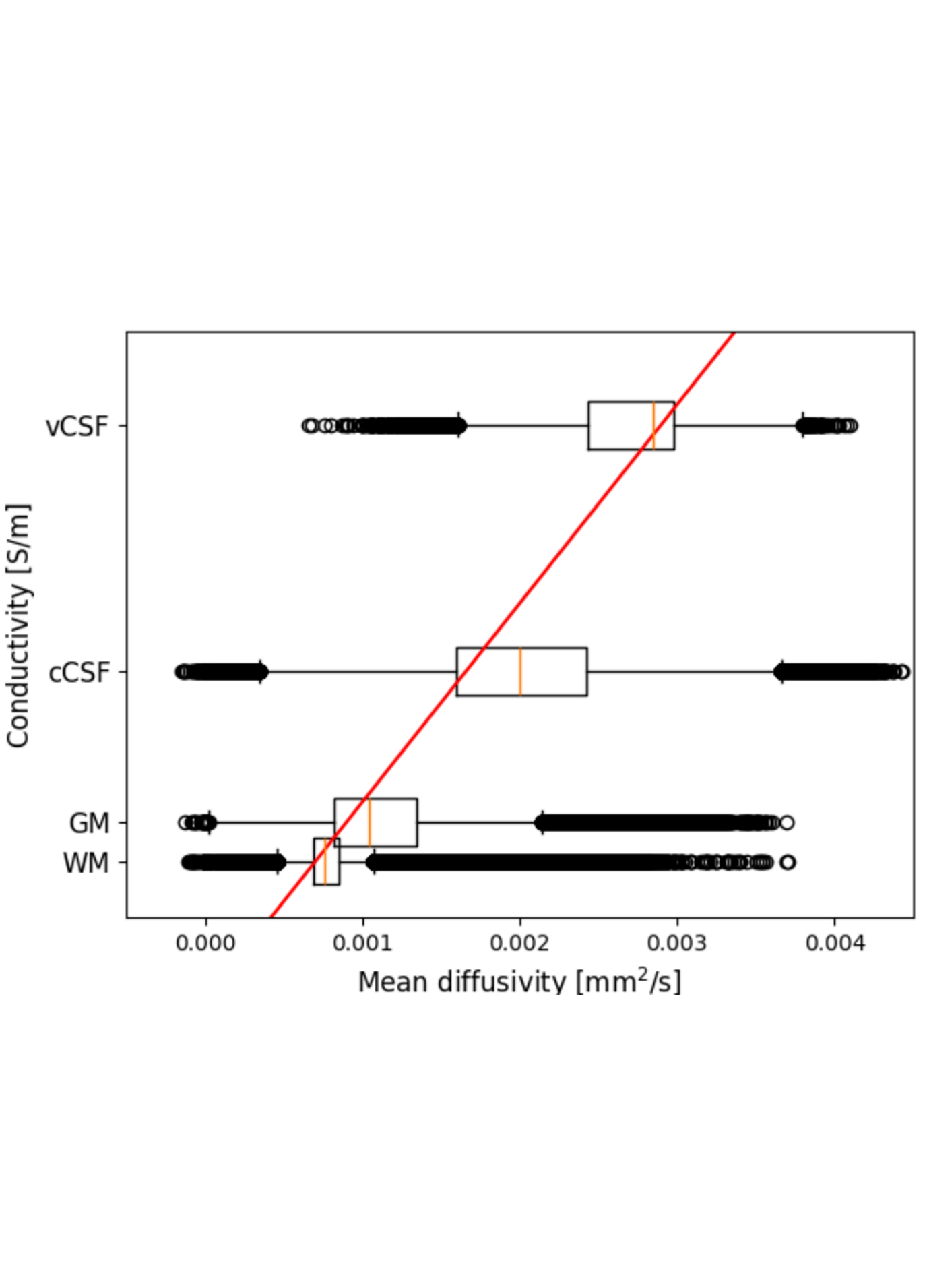

**Supplementary Figure 8. Linear fittings of tissue conductivities against the mean diffusivity values in a representative stroke patient.** Literature values of conductivities in ventricular cerebrospinal fluid (vCSF) and gray matter (GM) were fitted against the corresponding mean diffusivity values for each patient. cCSF: cortical CSF, WM: white matter

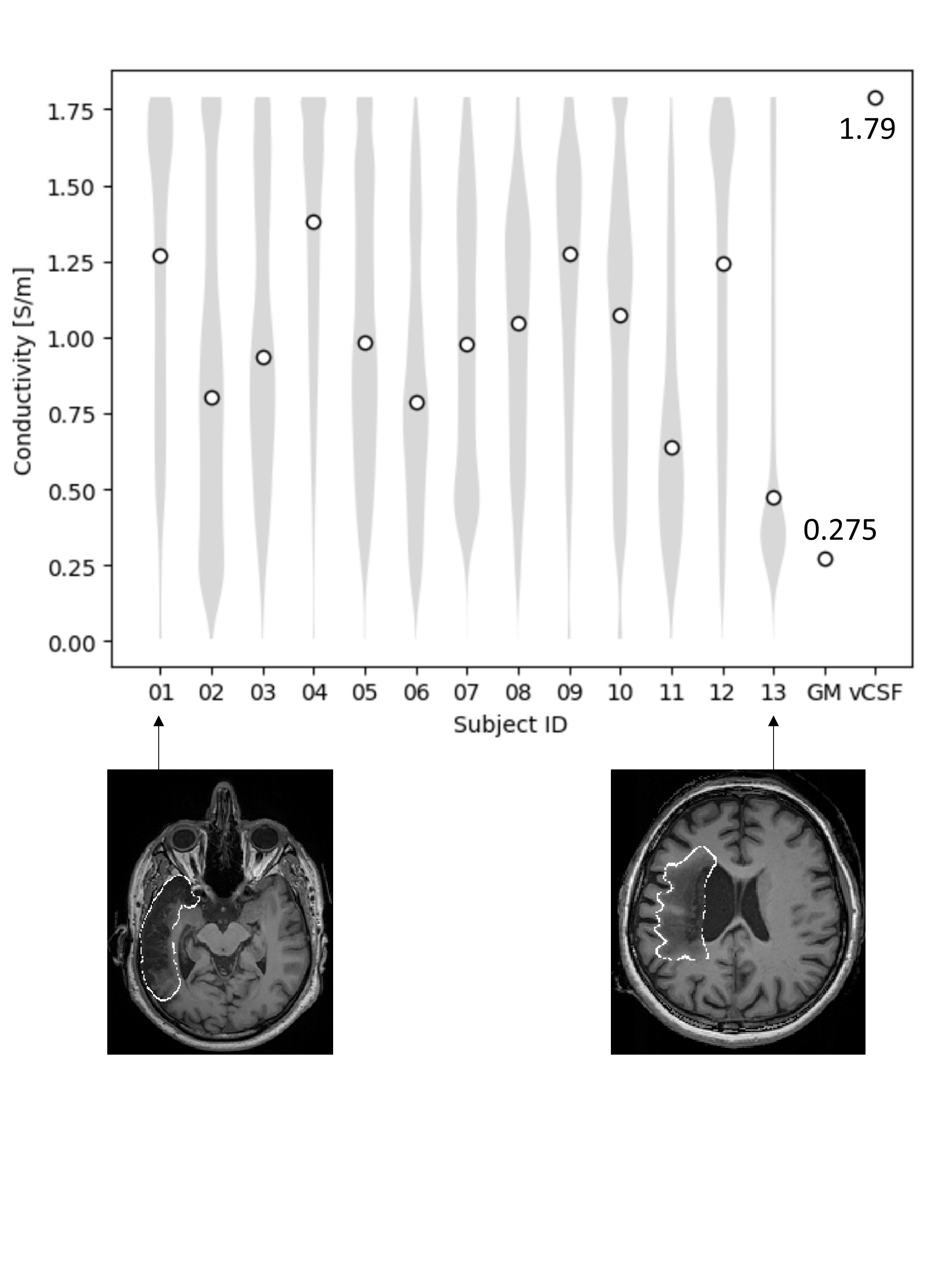

**Supplementary Figure 9. Estimated conductivities of lesions in stroke patients using the diffusion-to-conductivity mapping.** Each violin plot shows the estimated conductivities within the lesion masks for each stroke patient. The literature values of the ventricular cerebrospinal fluid (vCSF) and gray matter (GM) are also shown for reference. The estimated conductivities reflect the substantial intra- and inter-individual variability of tissues properties in lesions. The lower panels show the T1-weighted image of the representative stroke patients. The lesions are surrounded by white lines.

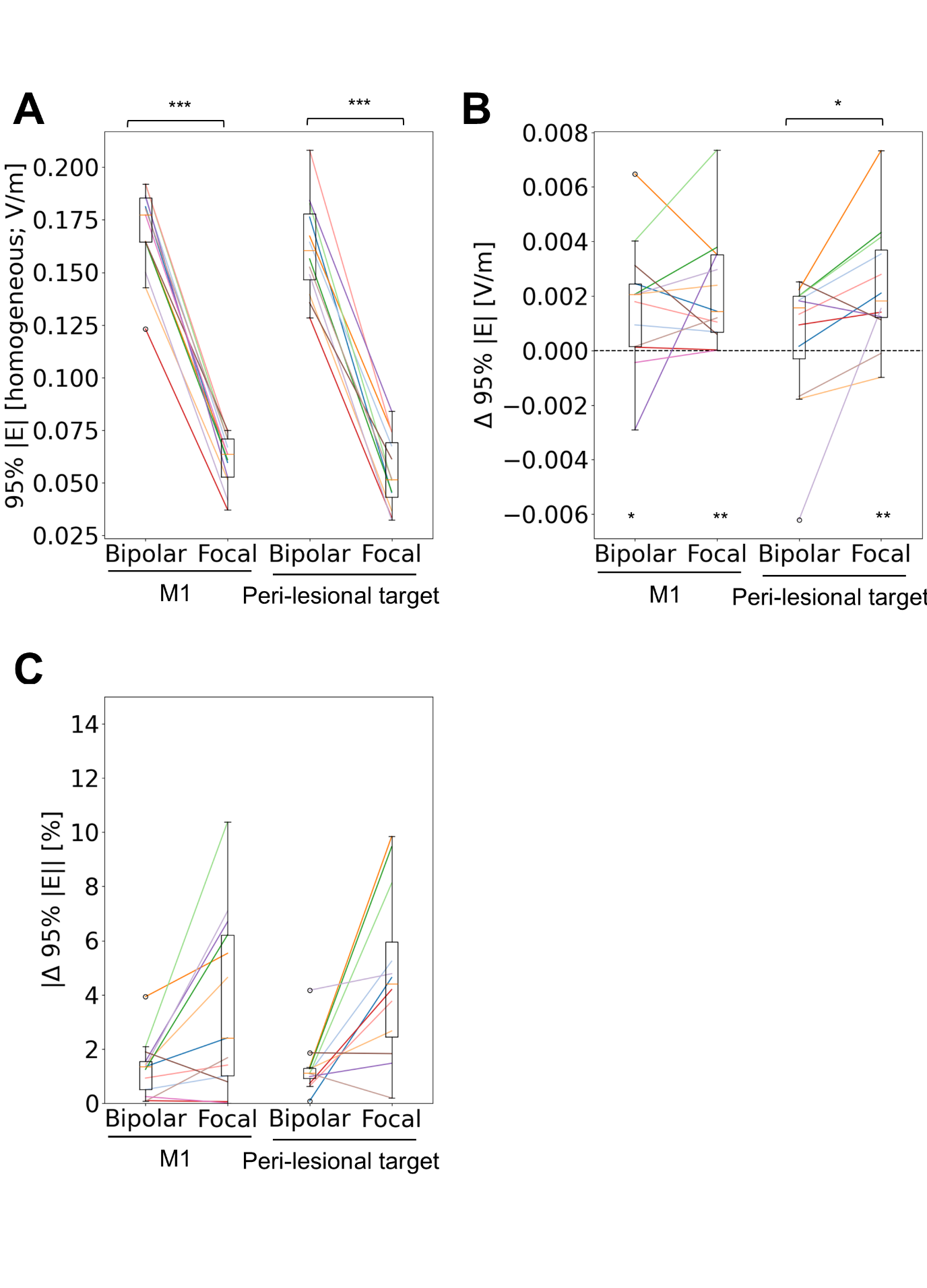
**Supplementary Figure 10. Change in the 95^th^ percentile peak magnitude of electric field (95% |E|) within the gray matter after changing lesion models from homogenous to realistic.** (A-C) Boxplots of 95% peak |E| in the homogeneous-lesion head models (A) and the change after changing to the realistic-lesion head models (B and C) for each montage (target location [the hand representation of left primary motor cortex (M1) or peri-lesional target] x type [bipolar or focal]). A and B show the actual value (V/m), while C indicates the relative absolute difference (%). * *P* < 0.05 ** *P* < 0.01 *** *P* < 0.001

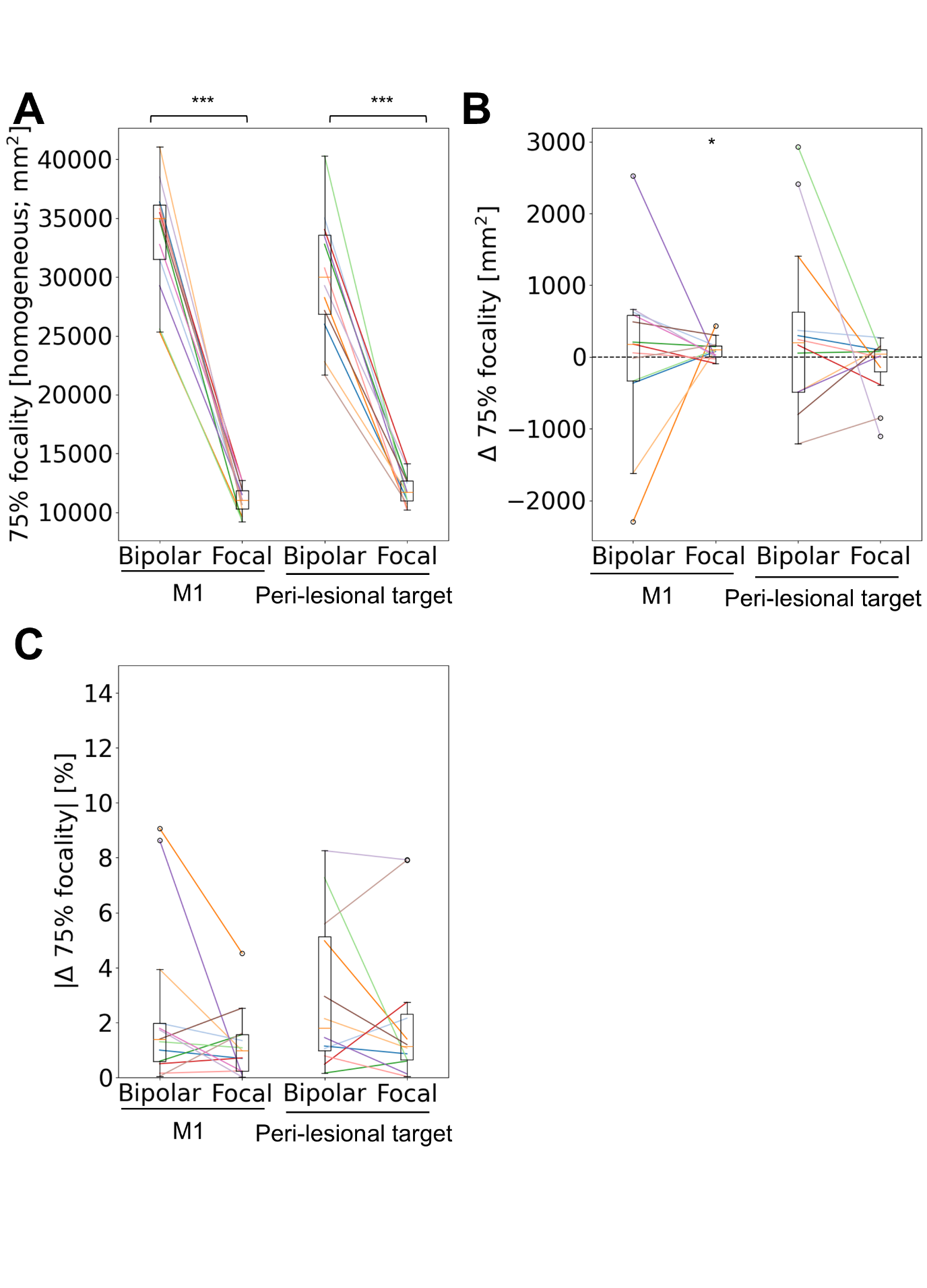

**Supplementary Figure 11. Change in the focality of electric field (E-field) within the gray matter after changing lesion models from homogenous to realistic.** Focality was defined as the area more than 75% above the 95^th^ percentile of the magnitude of E-field (75% focality). (A-C) Boxplots of 75% focality in the homogeneous-lesion head models (A) and the change after changing to the realistic-lesion head models (B and C) for each montage (target location [the hand representation of left primary motor cortex (M1) or peri-lesional target] x type [bipolar or focal]). A and B show the actual value (mm^2^), while C indicates the relative absolute difference (%). * *P* < 0.05 *** *P* < 0.001
